# Human–Agentic AI Collaboration Workflow for Trend Analysis of ChatGPT Applications in Healthcare

**DOI:** 10.64898/2026.11.14.26368120

**Authors:** Po Ju Chou, Yu Hsiu Lin, Eldon Y. Li

## Abstract

**Background:** With the rapid advancement of artificial intelligence (AI), large language models (LLMs) such as ChatGPT have demonstrated strong potential to transform healthcare research and practice. However, LLMs are probabilistic and generate inherently variable outputs, which create risk and uncertainty in healthcare applications. Therefore, analyzing the current research landscape of ChatGPT applications is imperative to understand applied techniques and related concerns and to facilitate their practical integration into healthcare information systems.

**Objective:** Leveraging LLM-based agentic AI with domain experts to analyze trends in ChatGPT healthcare applications and to examine the collaborative and interpretative capacities of LLM agents in broader contexts beyond narrowly defined topics.

**Methods:** The review used a systematic search of the PubMed and Web of Science databases, following PRISMA-ScR guidelines, and focused on ChatGPT applications, associated concerns, applied techniques, and usage factors. To support the review process, an adaptive, iterative LLM-based agentic AI with a human-in-the-loop mechanism was employed for automated metadata extraction, identification, classification, and knowledge synthesis. The extracted information was subsequently used for temporal and thematic analyses, citation network construction, and main-path analysis.

**Results:** Across 192 papers, the review identified 511 ChatGPT application types, 690 concern items, and 368 influence usage factors distinct from traditional IT, achieving a metadata error rate of 1.04%. The human-agentic AI workflow enabled efficient synthesis of contextual information across studies, addressing the growing challenge of timely evidence integration in medical knowledge. The findings provide insights into the evolving trajectories of ChatGPT applications in healthcare and demonstrate the value of an expert-validated human-agentic AI workflow in supporting literature analysis and streamlining review processes.

**Conclusion:** This review provides a comprehensive overview of emerging ChatGPT applications, associated concerns, applied techniques, and usage factors in healthcare. The study further provides meaningful insights into the operational mechanisms of agentic AI, demonstrating the potential of human–AI collaboration for medical informatics research.

## Introduction

In 2022, IBM and O’Reilly reported that about 30% of companies use AI, 40% are evaluating adoption, and 30% have no plans to use it [1, 2]. By 2025, the Artificial Intelligence Index Report indicated that global private investment in generative AI surged by 18.7% in 2024, while organizational adoption of AI jumped from 55% to 78% [3]. Today, with rapid technological evolution, AI applications in healthcare have become ubiquitous, driven by academic and corporate researchers [4]. Among the many cutting-edge AI technologies, ChatGPT has garnered significant attention for its ability to generate coherent, accurate responses that resemble human language and thought, leading to its widespread adoption in the medical field [5, 6, 7]. ChatGPT, a Generative AI (GenAI) model, was trained on a 570 GB dataset from open internet sources and has 175 billion parameters [8, 9, 10], enabling it to handle Natural Language Processing (NLP) tasks by predicting the next word based on learned patterns [5].

Acknowledging its potential, Microsoft Research, together with OpenAI, has been studying the possible uses of LLMs in medical applications, documentation, data interoperability, diagnosis, research, and education [9]. Moreover, as AI tools become increasingly accessible, more patients and medical professionals are turning to them for medical information [11]. Beyond these practical applications, the exponential growth of medical knowledge has created significant challenges for medical students and clinical teams, making it crucial for AI to supplement and enhance their knowledge to ensure the best possible patient care [12].

Driven by practical demands, the evolution of related LLM technology has progressed from preliminary zero-shot prompting to more structured few-shot paradigms that incorporate explicit task instructions and self-verification mechanisms to achieve more accurate responses. Prompt engineering techniques, such as the ReAct loop (thought–action–observation), have enhanced LLMs’ reasoning and interaction capabilities [13] and have emerged as a crucial aspect of effective LLM use for various NLP tasks [14,15].

Retrieval-Augmented Generation (RAG) enables LLMs to access up-to-date external knowledge, improving factual accuracy and reducing hallucinations. Results from using LLMs with RAG technology show that LLMs can be effective at a lower cost and can efficiently prevent hallucinations [16]. Furthermore, the introduction of function-calling and tool-use mechanisms has transformed LLMs into executable agents capable of performing deterministic, domain-specific tasks [17, 18, 19]. At the same time, foundation models have evolved into reasoning- and code-oriented architectures, significantly enhancing contextual comprehension, reasoning stability, and task adaptability [20, 21].

Building on foundation LLMs, agentic AI further integrates LLM agents, prompt engineering, RAG technologies, and reasoning enhancements into a workflow that supports multi-agent collaboration and coordinated autonomy. As a result, agentic AI can exhibit goal-directed behavior, dynamic adaptation, and self-improvement [22]. Consequently, it has been increasingly deployed in research automation and medical decision support [17]. Gridach *et al.* [18] investigated agentic AI for scientific discovery to address system reliability and automate literature reviews, emphasizing the importance of human-AI collaboration. In addition, agentic AI demonstrates adaptability, advanced decision-making, and self-sufficiency, enabling it to operate dynamically in evolving environments to pursue complex goals with minimal human intervention [19].

However, one major limitation of LLMs is their tendency to hallucinate false or irrelevant information [23]. As a result, it is difficult for users to determine whether the provided answers are grounded in appropriate facts [24]. Additionally, scholars suggested that several barriers hinder clinical use, including the lack of up-to-date medical information, failure to consider specific patient circumstances, and insufficient medical expertise and background to understand the complex relationships between conditions and treatments [7, 8].

Moreover, medical AI tools can become unreliable due to biases in the training data, both known and unknown, which may reflect societal inequities [25]. Furthermore, healthcare organizations that aim to implement ChatGPT would need comprehensive guidelines for handling patient data and implementing measures to secure patient privacy [15]. Additionally, AI models are often unable to trace the evidence line, making auditability and verification difficult [26]. Unlike traditional rule-based, deterministic information technologies, LLMs are probabilistic, data-driven, and inherently intangible (black-box) [27], thereby generating capricious, unpredictable outputs. As a result, scholars noted that whether AI-based technologies actually deliver improved outcomes and added value to healthcare remains unanswered [28]. However, despite these challenges, applying LLMs has significant potential to enhance multiple aspects of healthcare [29]. The coexistence of opportunities and challenges can lead to either stagnation or progress, warranting further investigation.

Given the numerous barriers and challenges, analyzing the current research landscape and summarizing major application trends are imperative for developing a comprehensive understanding of these technologies, facilitating practical applications, and guiding future research. Such reviews help identify knowledge gaps, enabling more systematic advancement of research [30]. Lai *et al.* noted that the rapid pace of medical progress and evidence turnover has increased the demand for timely evidence synthesis, and that literature reviews can integrate and evaluate existing research to guide clinical decisions [31]. However, literature review processes are predominantly manual and time-intensive, requiring substantial effort for article screening, eligibility assessment, and data extraction, which becomes more difficult when researchers examine multiple themes, such as applications, techniques, and concerns, simultaneously.

In contrast, LLMs can process large volumes of literature simultaneously while providing rapid, iterative feedback to researchers. This capability enables iterative refinement of the review, improving the efficiency and responsiveness of evidence synthesis within the review process. Beyond following the traditional literature review, this study incorporates LLM agents with advanced RAG technologies to generate evidence-driven data automatically from the newest medical papers and to achieve the following research objectives:

1. Leverage LLM-based agentic AI in collaboration with domain experts to identify and analyze trends in ChatGPT healthcare applications.
2. Examining the collaborative and interpretative capacities of LLM agents in responding to human queries and interacting with one another.

Through inter-agent interaction, learning, reflection, and collaboration with domain experts, the approach offers deeper insights into trends in ChatGPT healthcare applications and enhances the literature review process and outcomes. This advancement would ultimately inform evidence-based strategies for integrating LLM-based agentic AI into medical research and clinical practice for general purposes.

## Materials and Methods

This study integrates a review process with human-agentic AI collaboration, incorporating prompt engineering, RAG technology, and LLM agents to extract and identify metadata from medical journal articles. The metadata extracted from the studies included: authors, title, publication date, journal, abstract, purpose, keywords, citation, applications, concerns, and usage factors. The evidence-driven extracted data were used to cluster and classify ChatGPT applications, applied techniques, and associated concerns. Through multiple data analyses, including temporal, thematic, and main path analyses [32, 33, 34], we trace knowledge flows and reveal critical trends in healthcare ChatGPT applications. Fig. 1 illustrates how LLM agents extract, identify, classify, and format metadata through iterative refinement, yielding optimized results that enabled expert-led temporal, thematic, and main-path analyses.

**Fig. 1.**
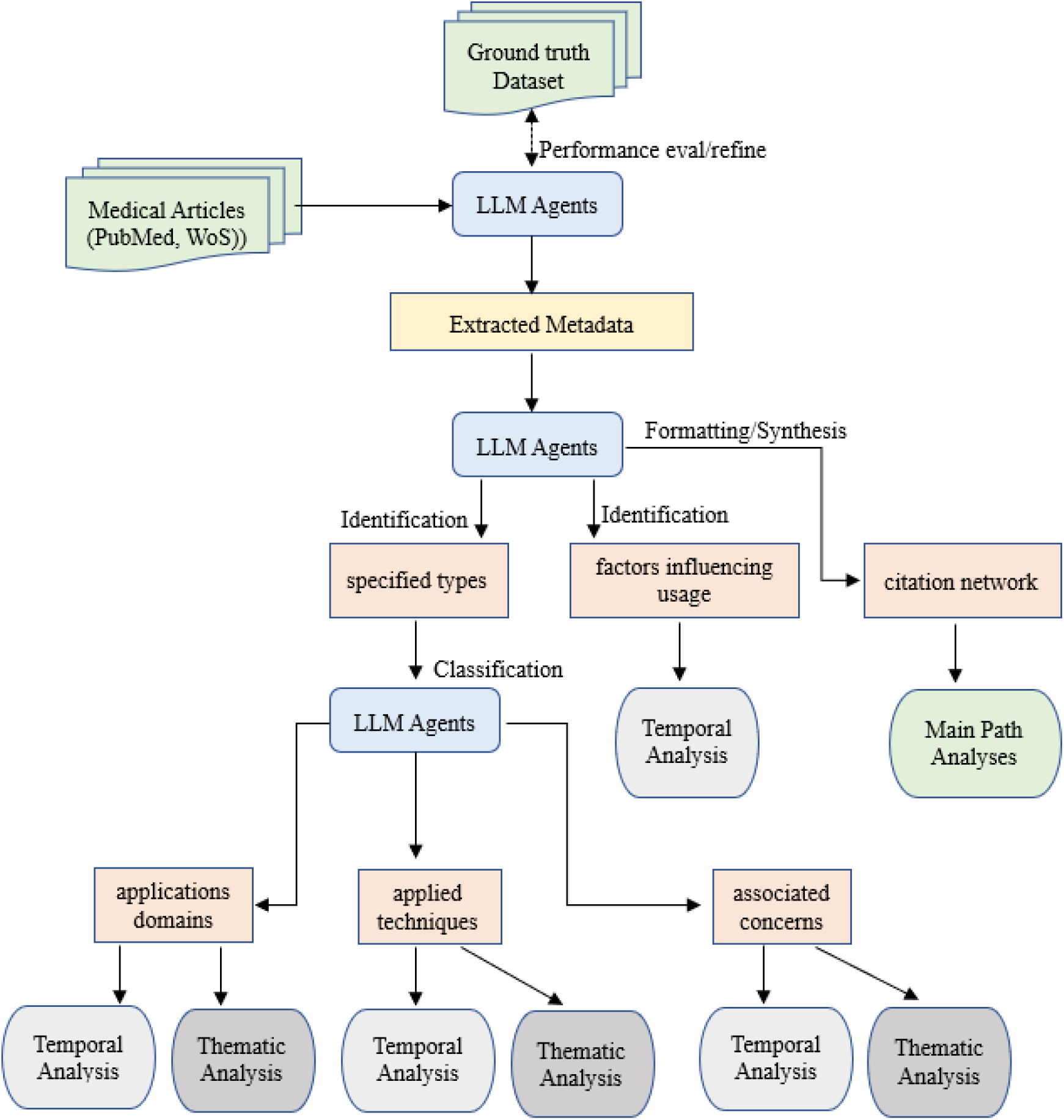
The metadata extraction/identification and multiple data analyses process

### Ground-truth Dataset

Initially, this study used 35 papers (see Supplementary file) from 16 medical journals as a ground-truth dataset to construct, evaluate, and iteratively refine prompting strategies, RAG retrievers, workflow component performance, and agent interaction, thereby optimizing agentic AI metadata extraction and identification results and reproducibility for subsequent multiple data analyses by domain experts.

### Data Collection

The keywords “ChatGPT,” “Healthcare,” “Medical,” “GenAI,” and “Agentic AI” were used to search the Web of Science (WoS) Core Collection and EndNote PubMed database. The search strategy was refined by publication year (2023–2025), open access status, and document type (article or review article).

### Criteria for Study Screening, Data Extraction and Synthesis

In this study, 2 reviewers conducted the screening. Only full-text articles were included in this scoping review to examine metadata related to ChatGPT applications, applied techniques, associated concerns, and usage factors. An agentic AI workflow was used to handle data extraction and validation automatically. Therefore, no further explicit inclusion or exclusion criteria were applied in this study. Some LLM agents were employed to extract, identify, reason, and synthesize data from the included papers, while other agents assessed generated responses using an LLM-as-a-judge approach for grading and verification. Within this iterative loop, human-AI cooperation was applied to optimize agent-generated responses. Different metadata extraction and synthesis forms were stored in separate CSV file formats. Finally, 3 reviewers validated the overall files and results.

### Human-Agentic AI Workflow

A goal-convergent spiral approach was implemented in this study, beginning with a small dataset and progressing from coarse structural schema to progressively expanded datasets and more detailed tasks toward the intended objective. Meanwhile, domain experts were integrated into the iterative human–AI collaborative agentic AI workflow to ensure accuracy and reproducibility. Fig. 2 illustrates how experts and LLM agents use the database and agentic RAG technology to leverage external data sources, retrieve relevant information, and validate generative responses. Throughout this pipeline, the human-in-the-loop approach ensures that data-driven discoveries are grounded in domain knowledge, validated appropriately, and translated into meaningful scientific advances rather than remaining isolated algorithmic outputs [35].

**Fig. 2.**
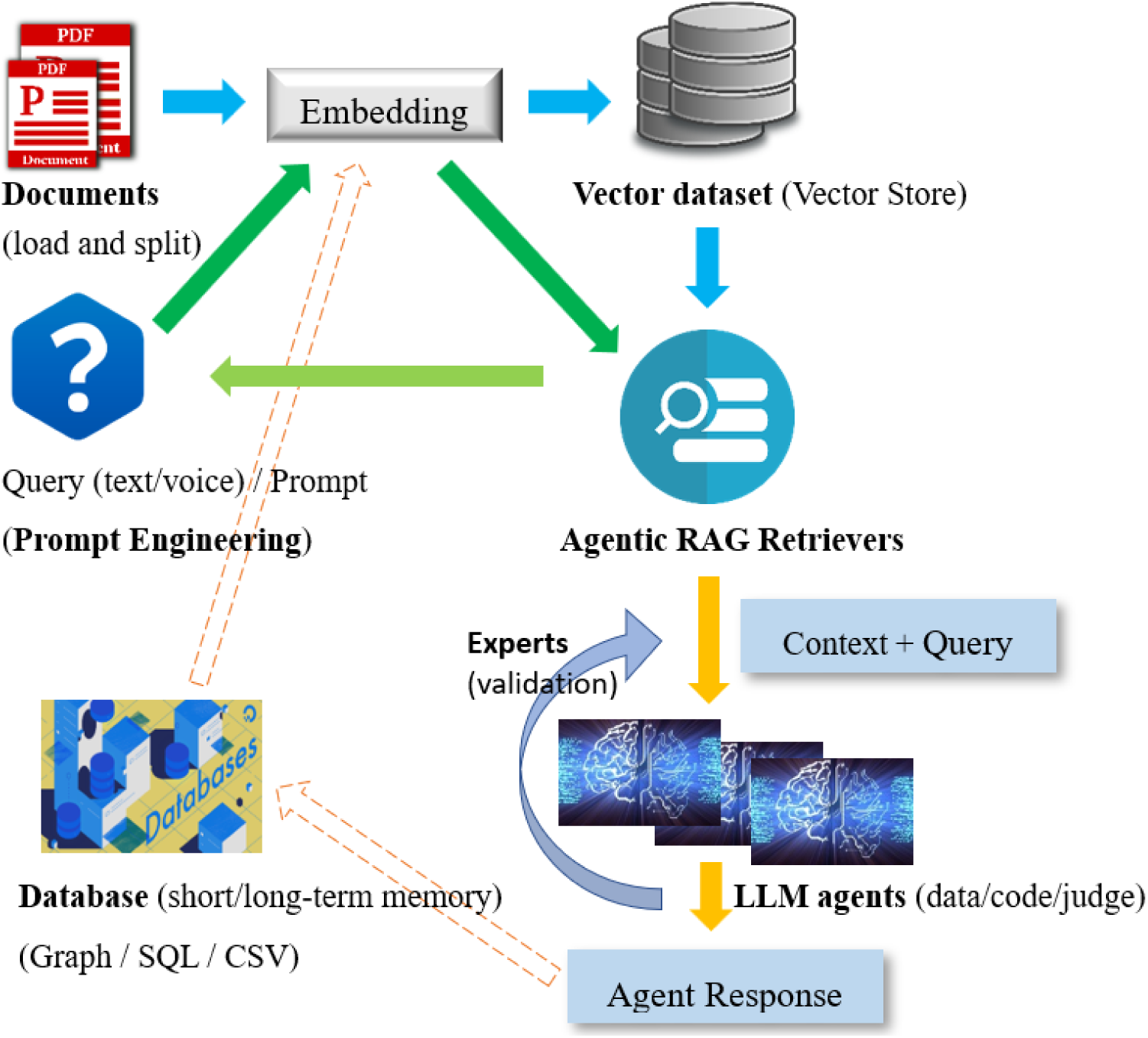
Pipeline of experts and LLM agents using Agentic RAG and databases The foundation models in Fig. 2 include Meta Llama 3.x, OpenAI turbo-GPT-3.5/GPT-4o-mini/GPT-5o, and a temperature setting of 0.0.

The proposed RAG-powered agentic AI consists of five major components (Loader, Converter/Embedding, Retrievers, LLM-based agents, Databases). Each component can be independently designed and validated before integration into a complete system. Generated outputs were evaluated and validated through a reflection process using code-based functional tools for analysis and visualization, as well as LLM-based agents for grading, to recheck and correct results. Validation also involved experts cross-checking LLM-generated outputs against source documents to ensure accuracy and contextual consistency. Through prompt engineering and iterative refinement, the optimal configuration of the components and the overall pipeline can be identified.

## Multiple Data Analyses

### Temporal Analysis and Thematic Analysis

Seed articles were stratified into six-month intervals to examine temporal trends in ChatGPT across applications, applied techniques, and associated concerns. LLM agents processed the seed articles to generate terms for classifying applications and associated concerns, while applied techniques were classified directly into five categories from the articles. Thematic analyses were conducted on the data generated by LLM agents to establish distinct taxonomies for ChatGPT applications, applied techniques, and associated concerns.

### Factors Influencing Usage

The LLM agents extracted and evaluated the text spans describing ChatGPT adoption or usage conditions based on GenAI systems’ unique features. These spans identified specific factors to distinguish GenAI adoption and ongoing use from conventional IT implementation in healthcare.

### Citation Networks and Main Path Analysis

This study constructed a citation network to examine the evolution of ChatGPT application research and applied Main Path Analysis (MPA) to identify the principal knowledge trajectories within the network, with a specific focus on the Global Main Path (GMP) [34]. We extracted the reference lists from each of the 192 seed papers using GROBID (GeneRation Of Bibliographic Data), then parsed and converted them to APA format using an LLM agent to construct a citation network. To compute the GMP, we used the Search Path Count (SPC) approach, which quantifies each document’s (node) importance by counting the number of citation paths that traverse it, thereby identifying the most influential end-to-end path [34].

## Results

A total of 60 full-text articles were retrieved from the Web of Science and 104 articles from PubMed. After excluding 7 duplicates, the remaining 157 were integrated with the 35 ground-truth articles, forming a final corpus of 192 seed papers (see Fig. 3).

**Fig. 3.**
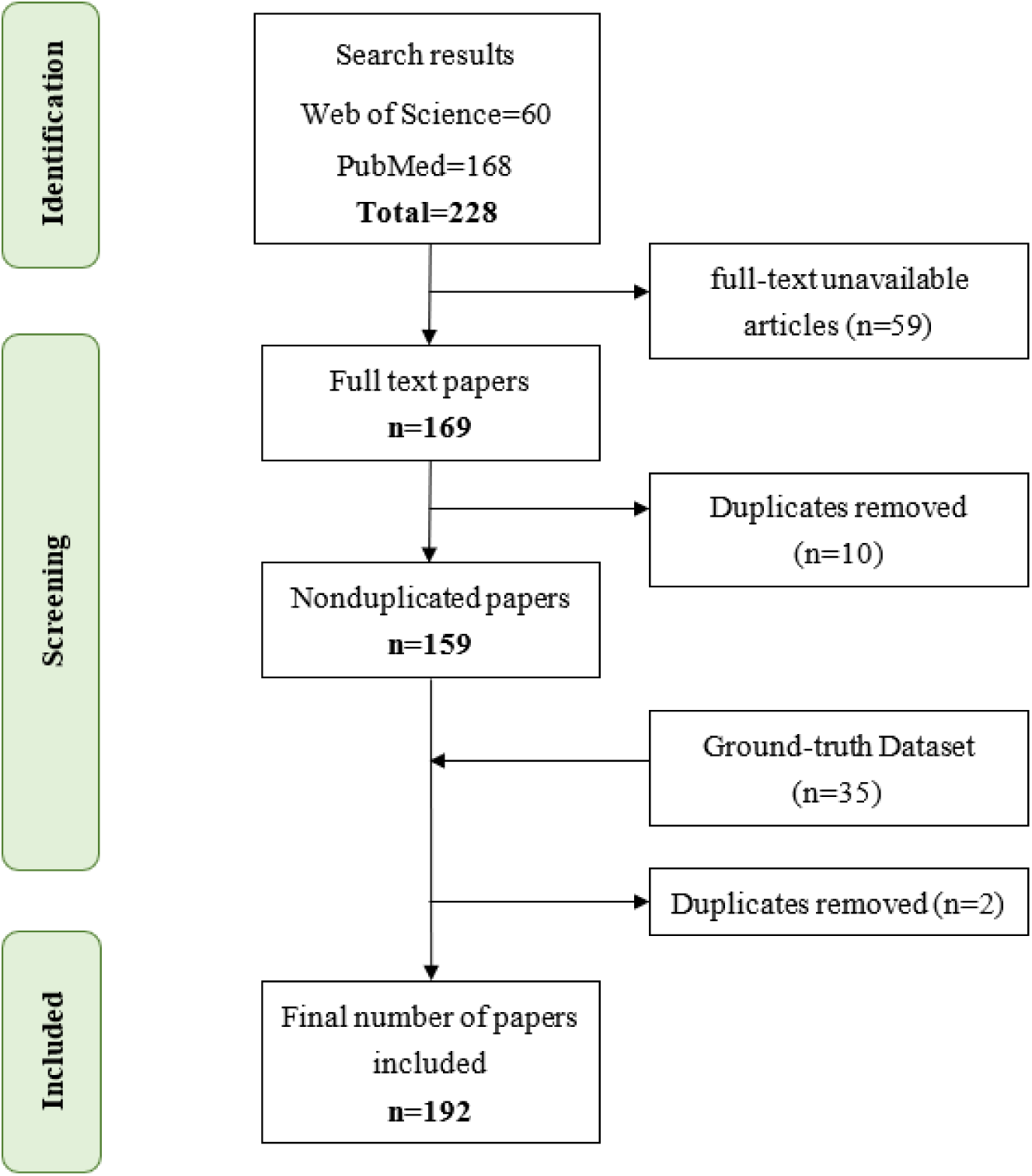
Flow diagram of the studies screening and selection

Following numerous iterations across 192 articles, metadata fields, and repeated test runs from the initial prototype to the final workflow framework, the LLM agents correctly extracted and identified metadata such as title, authors, abstract, journal, keywords, and paper citations. Of 192 articles. 1 article lacked the published date, and 2 (1.04%) were misidentified. ChatGPT applications were not identified in 1 article (0.5%), as associated concerns were not identified in 2 articles (1.04%), and factors influencing usage were not identified in 2 articles (1%). After the reviewers examined the final results, the authors corrected all errors and omissions and confirmed them.

### Key Characteristics of the Seed Papers

As shown in Table 1, each year is divided into two 6-month periods, H1 and H2. H1 covers January to June, and H2 covers July to December. The number of reviewed seed articles across periods increased from 29 in 2023-H1 to a peak of 68 in 2024-H1, reflecting a growing research trend, before declining sharply from 2024-H2 onward. Among 192 articles, 117 journals were identified as relevant. *Cureus* published the most articles (16), followed by *npj Digital Medicine* (6), *Healthcare* (5), *PLOS Digital Health* (4), and *Frontiers in Artificial Intelligence* (3). Additionally, 34 journals published 2 articles each, while 78 published one article each.

**Table 1.** Key characteristics of 192 seed articles.

| Properties | Category | count | % |
|---|---|---|---|
| Number of articles published (192 total) | 2023-H1 | 29 | 15.1 |
|  | 2023-H2 | 47 | 24.5 |
|  | 2024-H1 | 68 | 35.4 |
|  | 2024-H2 | 18 | 9.4 |
|  | 2025-H1 | 18 | 9.4 |
|  | 2025-H2 | 12 | 6.2 |
| Top 5 Journals<br>in WoS and PubMed | Cureus | 16 | 8.3 |
|  | npj Digital Medicine | 6 | 3.1 |
|  | Healthcare | 5 | 2.6 |
|  | PLOS Digital Health | 4 | 2.1 |
|  | Frontiers in Artificial Intelligence | 3 | 1.6 |
| ChatGPT Applications | Clinical Decision Support | 181 | 94.3 |
|  | Education (Medical and Health) | 112 | 58.3 |
|  | Medical Research and Data Analysis | 110 | 57.3 |
|  | Healthcare/Medical Workflow | 75 | 39.1 |
|  | Healthcare/Medical Consultation | 69 | 35.9 |
|  | Medical Documentation | 66 | 34.4 |
|  | Public Health | 7 | 3.6 |
| Applied Techniques | Retrieval-Augmented Generation | 98 | 51.0 |
|  | Prompt Engineering | 86 | 44.8 |
|  | Agent and Multi-Agent | 48 | 25.0 |
|  | Agentic AI | 27 | 14.1 |
|  | Fine-Tuning | 14 | 7.3 |
| Associated Concerns | Ethical, Safety, and Legal Concerns | 177 | 92.2 |
|  | Reliability, Explainability, and Interpretability Challenges | 160 | 83.3 |
|  | Collaboration and Trust | 100 | 52.1 |
|  | Implementation and Knowledge-Management Barriers | 98 | 51.0 |
|  | Data and Knowledge Limitations | 98 | 51.0 |
|  | Evaluation and Validation Deficiencies | 78 | 40.6 |

Most ChatGPT application studies pertained to Clinical Decision Support (181 articles), followed by Education (112), Medical Research and Data Analysis (110), Healthcare/Medical Workflow (75), Healthcare/Medical Consultation (69), Medical Documentation (66), and Public Health (7). Regarding applied techniques, RAG accounted for 98 studies, followed by Prompt Engineering (86), Agent and Multi-agent approaches (48), Agentic AI (27), and Fine-Tuning (14).

Ethical, Safety, and Legal Concerns accounted for the highest proportion with 177 articles, followed by Reliability, Explainability (160), and Interpretability Challenges (160), Collaboration and Trust (100), Implementation and Knowledge Management Barriers (98), Data and Knowledge Limitations (98), and Evaluation and Validation Deficiencies (78).

### Specific application types within Application categories

The LLM agent extracted 511 specific application types in 7 major application categories from the 192 articles. Table 2 summarizes the frequency of reported specific application types in each application category. The 192 articles discussed 871 applications in total; each article may discuss one or more applications. Overall, the distribution was highly skewed toward the Clinical Decision Support category, with the majority of applications (155) in that category. Within the Education (Medical and Health) category, clinical education (40) and medical education (22) were most common, followed by patient education (15). The Medical Research and Data Analysis category is led by medical research (55). Applications in the Medical Documentation category were less prevalent overall, with medical Writing (8) and medical documentation (4) most prevalent. Likewise, the Healthcare/Medical Consultation and Healthcare/Medical Workflow categories were sparsely represented, with most applications focusing on patient communication (5). Finally, the Public Health category is the least prevalent, with healthcare information dissemination as the leading application (n=2).

**Table 2.**
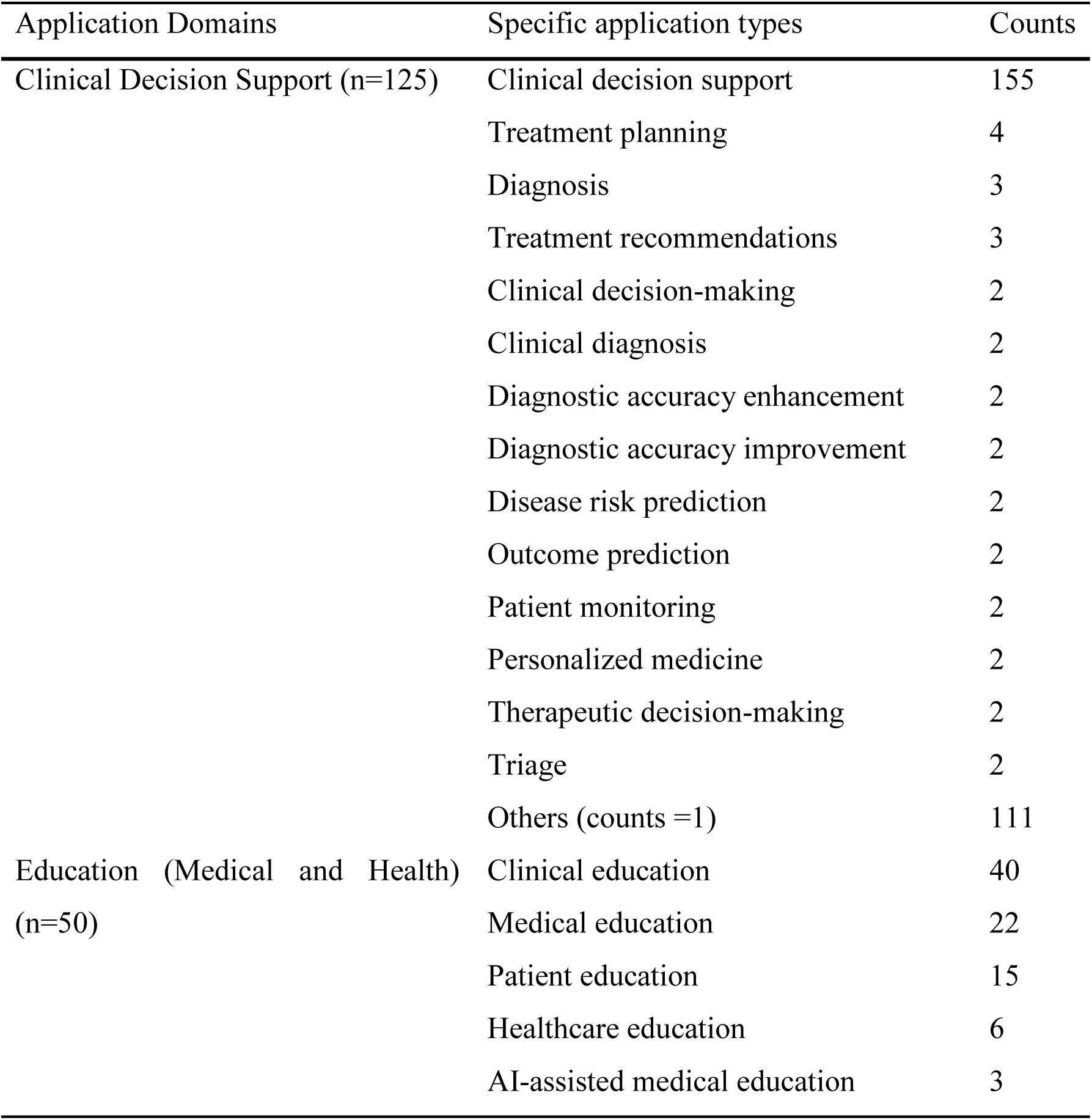
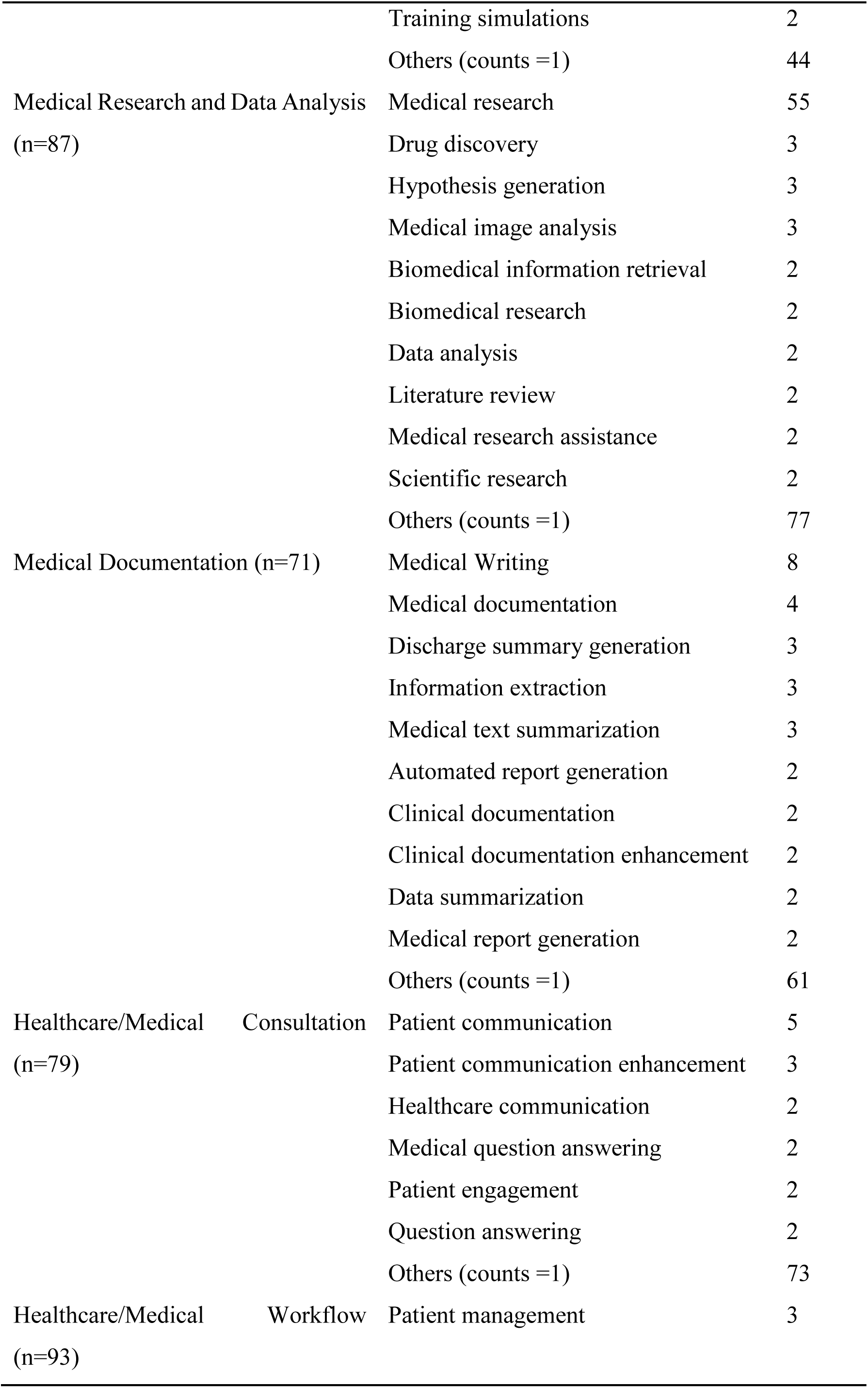
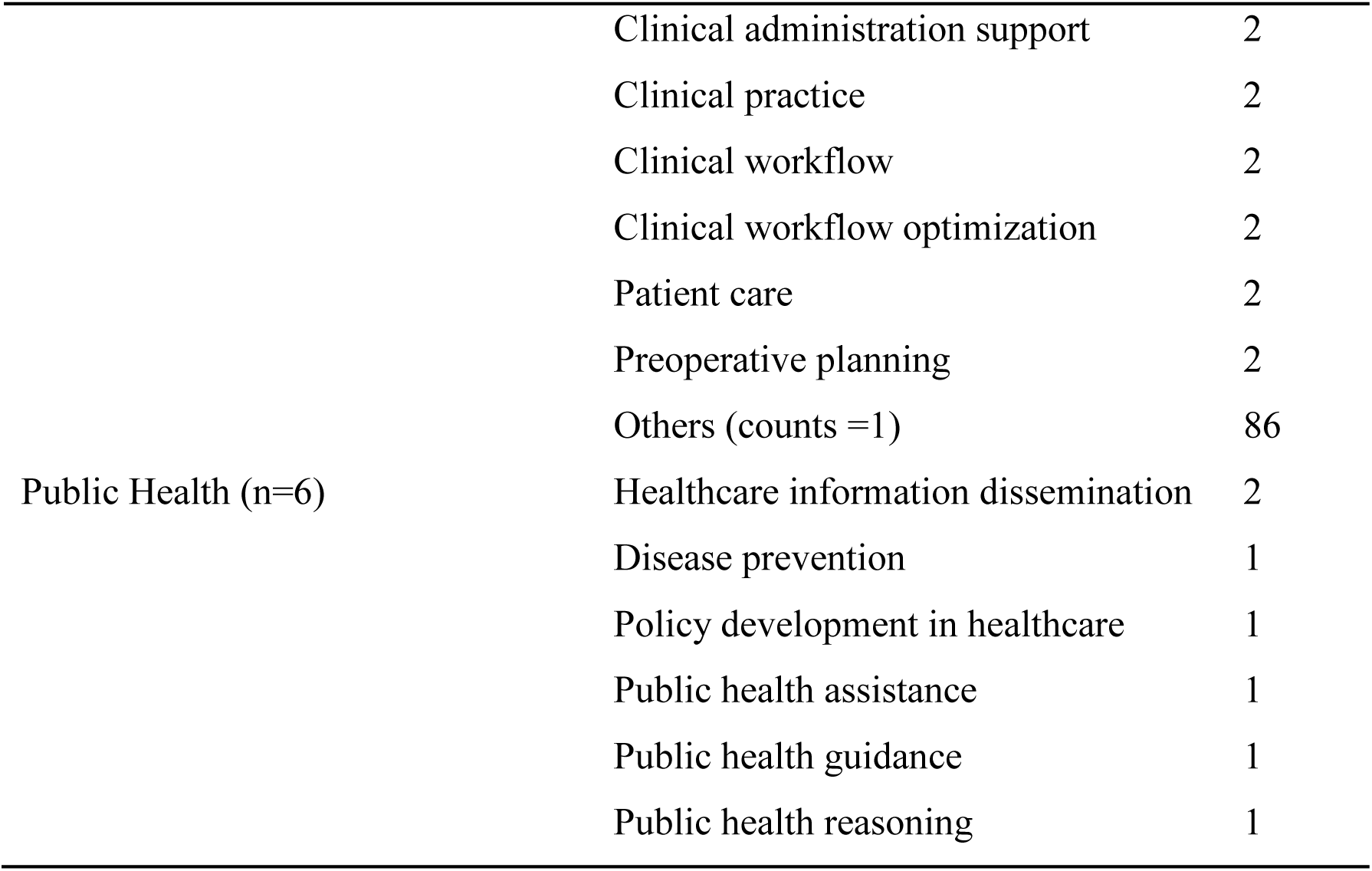
Distribution of reported specific application types across major categories (n=511).

### Specific Concern Types within Major Associated Concerns

A total of 690 specific concern types in 6 associated concern categories were identified from the seed papers. Among the major categories, Ethical, Safety, and Legal Concerns accounted for the highest number of reported issues (269), followed by Reliability, Explainability, and Interpretability Challenges (132), Implementation and Knowledge-Management Barriers (109), Data and Knowledge Limitations (81), Evaluation and Validation Deficiencies (66), and Collaboration and Trust (33). This distribution highlights the breadth of issues represented across the major concern categories.

Fig. 4 depicts the frequency with which specific concern types were identified within each overarching concern category (inclusion criterion: frequency ≥ 10). Within Ethical, Safety, and Legal Concerns, data protection (87) and ethical concerns (83) were the most prevalent, followed by bias (72), responsibility (68), data privacy (53), and patient safety (39). In Collaboration and Trust, trust emerged as a salient issue (76), whereas human oversight was reported less frequently (9). In the Reliability, Explainability, and Interpretability Challenges category, accuracy (70) and data accuracy (32) were the most prominent, followed by transparency (29), with the remaining specific concern types appearing at comparatively modest frequencies. In Data and Knowledge Limitations, data integrity (18) and data quality (13) were the most recurrent items. For Evaluation and Validation Deficiencies and Implementation and Knowledge Management Barriers, clinical validation (14) and data accessibility (11) were the most frequently reported, respectively.

**Fig. 4.**
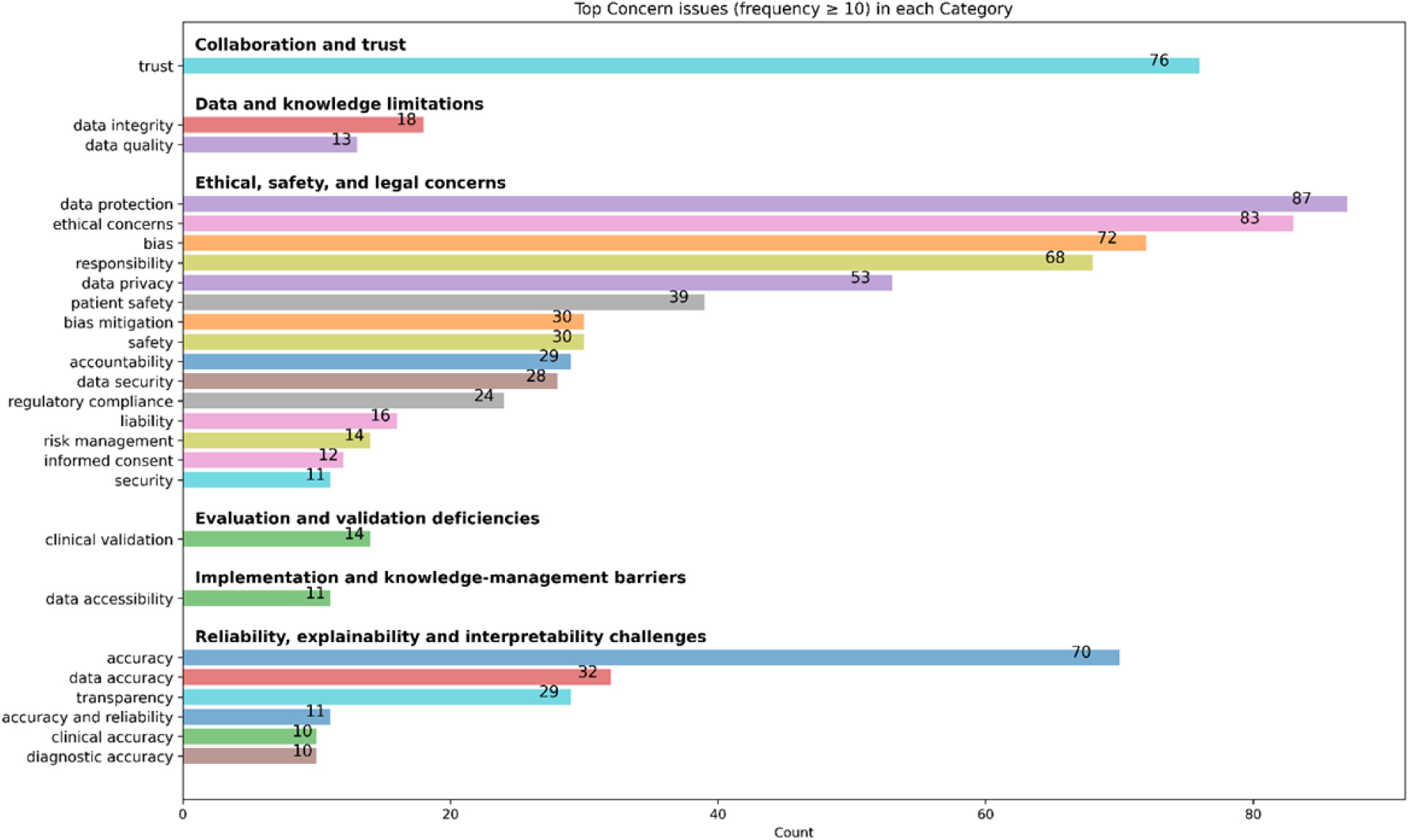
Frequency of specific concern types within major associated concerns

### Trend in ChatGPT Applications

Fig. 5-7 illustrate the temporal trends in ChatGPT applications, applied techniques, and associated concerns from 2023-H1 to 2025-H2. The numbers inside the circles denote the absolute count of studies, and the circle sizes correspond to normalized proportions relative to the total number of studies during the respective period. The values (n) displayed along the X- and Y-axes indicate the total number of studies.

**Fig. 5.**
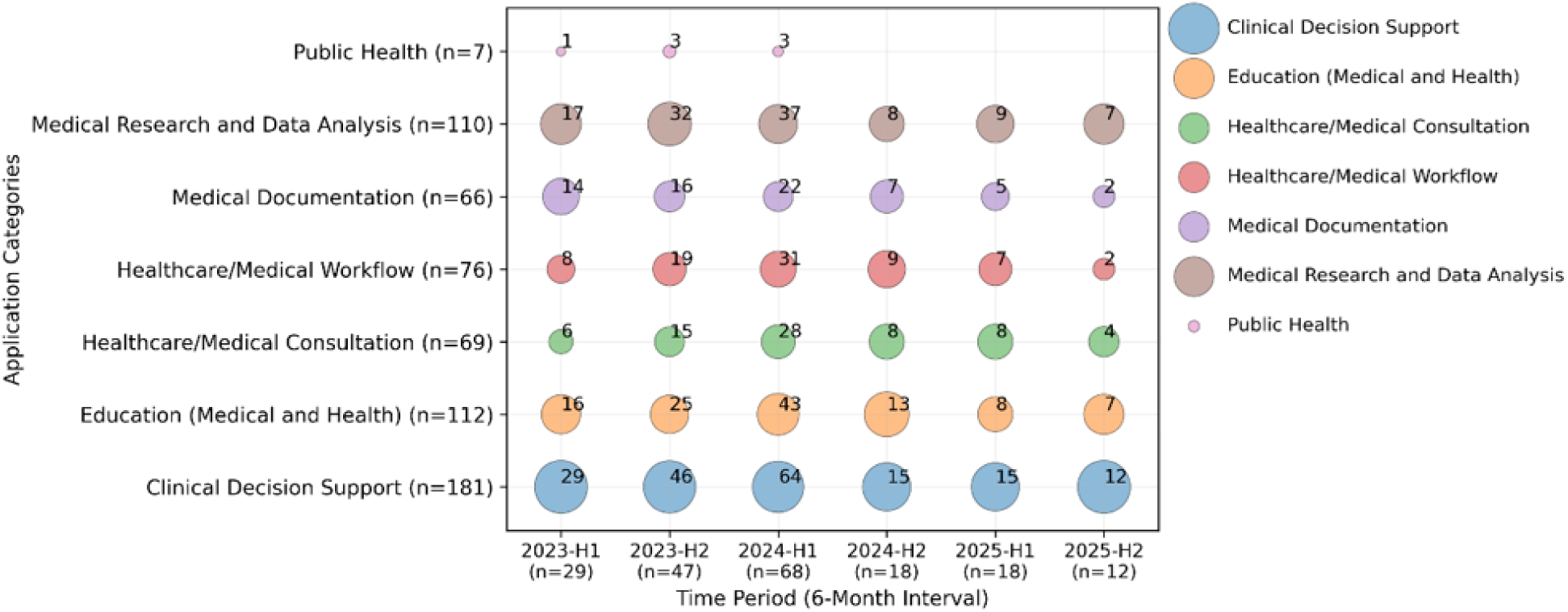
ChatGPT applications over time.

**Fig. 6.**
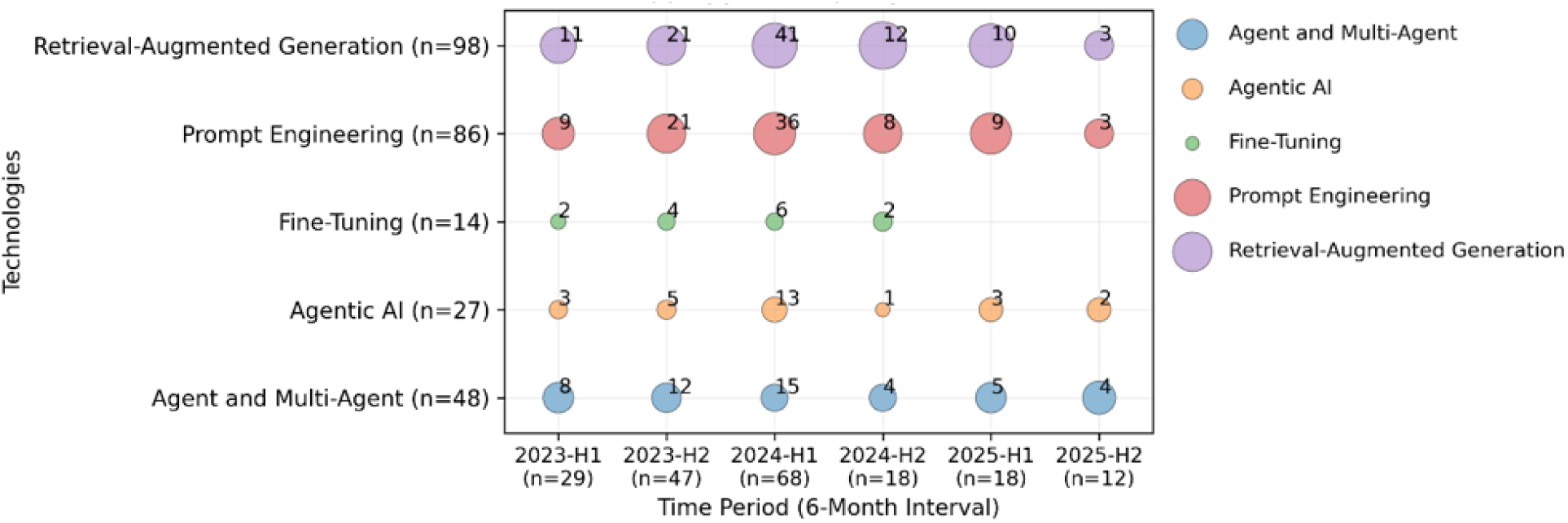
Applied techniques over time.

**Fig. 7.**
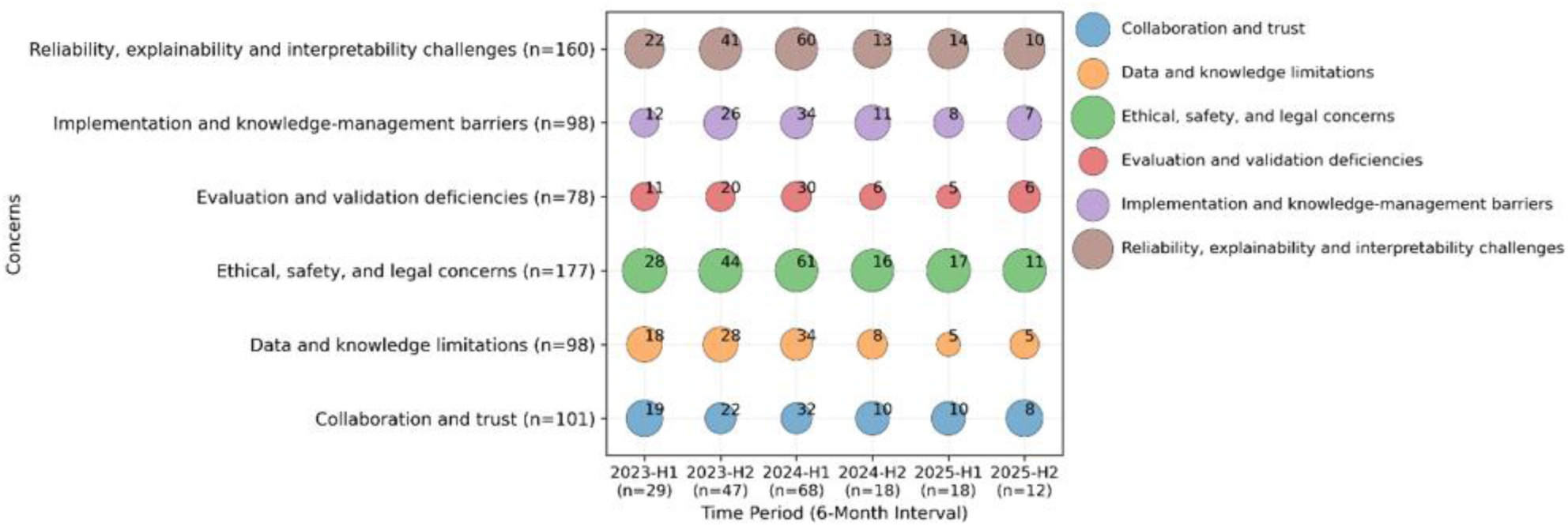
Concerns about ChatGPT applications over time.

As shown in Fig. 5, across all periods, Clinical Decision Support consistently accounted for the largest percentage of studies, followed by Education (medical and health) and Medical research and Data analysis, both of which peaked in 2024-H2 before declining. Healthcare/Medical Consultation, Healthcare/Medical Workflow, and Medical Documentation experienced substantial decreases in absolute counts and normalized proportions over time, while Public Health applications remained consistently low.

Of the 192 included articles, LLM agents identified 122 papers as related to the applied technique categories (see Supplementary file). Fig. 6 indicates the relative prominence of each applied technique category over time. RAG consistently showed the highest absolute counts and largest normalized representation across all periods, peaking in 2024-H1, followed by Prompt Engineering, which also demonstrated substantial but declining prominence after 2024-H1. In contrast, Agent and Multi-agent systems, as well as Agentic AI, exhibited an increasing trend after 2024-H2. Fine-tuning showed lower absolute counts and smaller normalized proportions over time, declining to zero after 2024-H2.

Fig. 7 presents temporal patterns in reported concerns about ChatGPT applications. Across all periods, Ethical, Safety, and Legal Concerns, as well as Reliability, Explainability, and Interpretability Challenges, were most frequently reported. Other concerns, including Evaluation and Validation Deficiencies, Implementation and Knowledge-management Barriers, Data and Knowledge limitations, and Collaboration and Trust, were reported less frequently. Overall, the concerns associated with each period exhibited relatively stable proportional relationships. Data and Knowledge limitations demonstrated a slight, gradual decline over time, whereas Evaluation and Validation Deficiencies and Collaboration and Trust showed a modest upward trend after 2024-H2.

A total of 368 factors that influence ChatGPT usage were extracted from 192 papers. They are distinct from traditional IT. Fig. 8 shows the top 10 most frequently reported factors: hallucination (101), output variability (93), trust issues (86), and interpretability (84). Additional high-frequency factors included context dependence (64) and transparency (54), followed by workflow integration challenges (43) and bias (41). Less frequent but still ranked among the top 10 were uncertainty (28) and non-deterministic behavior (15). Collectively, the dominant factors centered on reliability-related risks (e.g., hallucination and variability), explainability (interpretability and transparency), and socio-technical considerations (trust, bias, and workflow integration) affect adoption.

**Fig. 8.**
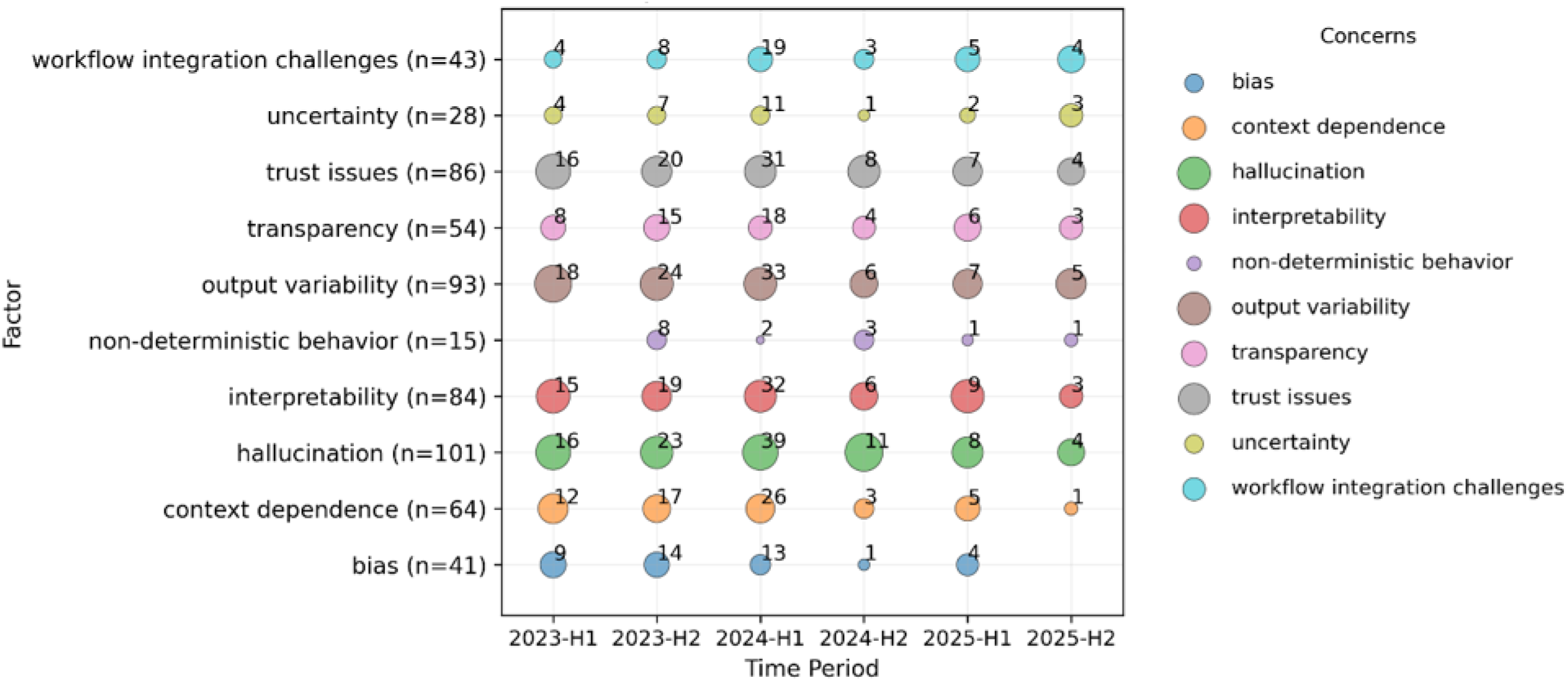
Top 10 factors influencing ChatGPT usage.

Fig. 8 illustrates that the distribution of the top 10 factors varied over time across 192 papers. Prior to 2024-H1, studies primarily focused on hallucination, output variability, trust issues, interpretability, transparency, context dependence, and bias. Workflow integration challenges, uncertainty, and non-deterministic behavior were more frequently observed during 2023-H2 and 2024-H1, whereas these issues were less prevalent or not prominently reported in other periods. After 2024-H2, hallucination, output variability, trust issues, interpretability, transparency, and workflow integration challenges remained prominent (average count >3), whereas the other 4 factors (context dependence, bias, non-deterministic behavior, and uncertainty) no longer appeared important. From 2025 onward, this study observed that several factors not included in Fig. 8 began to appear, including autonomy (4 in 2025-H1 and 3 in 2025-H2), emergent behavior (4 in 2025-H1), and ambiguity (3 in 2025-H1).

### Citation Network

After extracting the reference lists from the 192 seed articles, LLM agents identified approximately 6,206 unique articles across 8,962 referenced publications, along with associated metadata (title, authors, publication year, journal, DOI, citation counts, and in-text citations). From this data, we constructed 8,770 citation paths (edges) and identified 3,392 unique journals. Table 3 presents the characteristics of the publications in the citation network.

**Table 3.** Key characteristics of the citation network.

| Item | Article/ Journal/Sub-network/Year | n |
|---|---|---|
| Unique Papers |  | 6,206 |
| Unique Journals |  | 3,392 |
| Citation edges | whole citation network | 8,770 |
|  | connection path of 192 seed articles | 475 |
| Number of papers cited (2020-2024) | Year 2020 | 409 |
|  | Year 2021 | 526 |
|  | Year 2022 | 615 |
|  | Year 2023 | 2113 |
|  | Year 2024 | 959 |
| Top Journals | arXiv, arXiv preprint | 528, 43 |
|  | Cureus | 231 |
|  | medRxiv | 117 |
|  | Nature | 102 |
|  | JAMA | 91 |
|  | Radiology | 76 |
|  | Journal of Medical Internet Research, J Med Internet Res | 73, 67 |
|  | Healthcare | 72 |
| Top 10 cited papers | Kung et al, 2023 [39] | 46 |
|  | Sallam, 2023 [40] | 36 |
|  | Drazen et al, 2023 | 30 |
|  | Singhal et al, 2023 [37] | 29 |
|  | Cascella et al, 2023 [8] | 25 |

The citation network, derived from seed articles and their references, shows that cited references steeply peaked in 2023, whereas seed articles peaked in the first half of 2024. These temporal patterns suggest heightened scholarly attention during 2023–H2 and 2024–H1. However, unexpectedly, the number of reviewed publications declined sharply after that. The discrepancy may reflect publication or indexing lags, or a shift in research focus, and should be re-examined as more recent data become available.

### Main Path Analysis of Knowledge Flow

In Fig. 9, the Main Path Analysis (MPA) illustrates the knowledge flow of LLMs applications in medical research. The knowledge flow reveals a developmental trend from exploratory use and task-level capability validation to risk consolidation and ethical–educational integration, with concerns increasingly outweighing gains in technical maturity. The MPA identifies influential studies by tracing frequently traversed citation paths from the seed papers. The upstream segment of the main path is characterized by studies evaluating LLM performance on medical tasks, with explicit attention to accuracy, reasoning quality, and potential clinical harm. Singhal *et al.* [37] assessed LLMs through standardized medical benchmarks and human evaluations. Closely connected, Jeblick *et al.* [38] reported generally acceptable output quality alongside recurrent errors and safety-related issues.

**Fig. 9.**
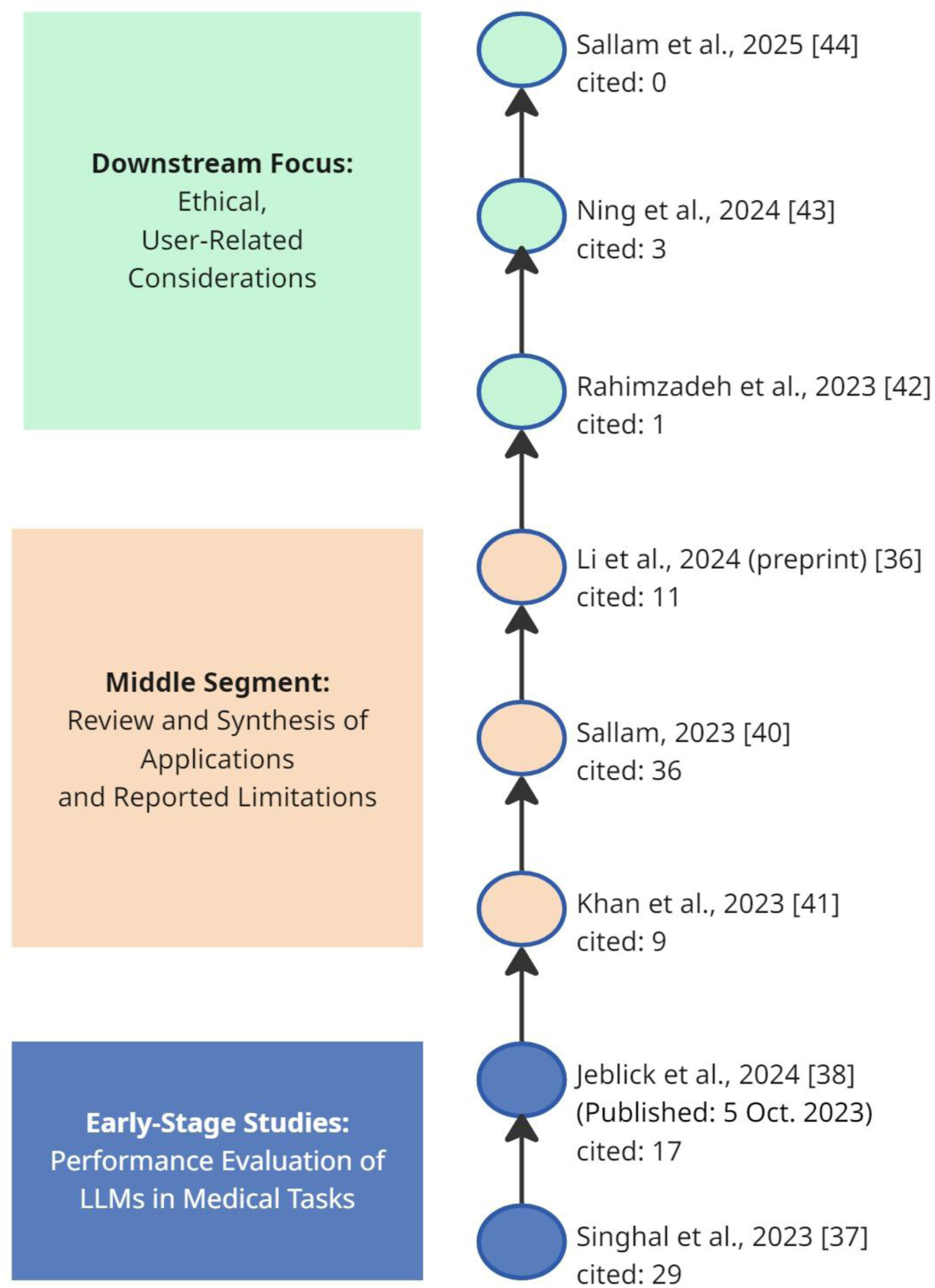
Knowledge flow along the main path of ChatGPT healthcare applications.

The middle segment of the main path is dominated by review-based studies that aggregated reported applications and limitations across medical domains. Analyses by Li *et al.* [36], Sallam [40], and Khan *et al*. [41] synthesized evidence on the use of LLMs in healthcare education, research, and clinical workflows. Across these reviews, recurring concerns included hallucination, bias, lack of transparency, ethical and legal uncertainty, and insufficient reliability for unsupervised clinical use, with limited reporting of domain-specific model deployment in clinical practice.

Downstream studies along the main path focused on ethical considerations and user-related responses to generative AI in healthcare settings. Rahimzadeh *et al.* [42] examined the use of ChatGPT as a supplementary tool in ethics education, while Ning *et al.* [43] proposed structured approaches for assessing ethical issues in GenAI research and applications. Sallam *et al.* [44] further contributed empirical evidence by measuring fear, mistrust, and ethical concerns among health sciences students.

### Knowledge Graph

A Neo4j graph database was constructed to store the extracted metadata and citation network, allowing papers and their relationships to be easily added, updated, searched, and visualized as required. Fig. 10 illustrates the citation context for Li et al. [36], which shows 6 incoming citations and 11 cited papers, with all 18 nodes connected and annotated by publication year, distributed across 17 distinct journals.

**Fig. 10.**
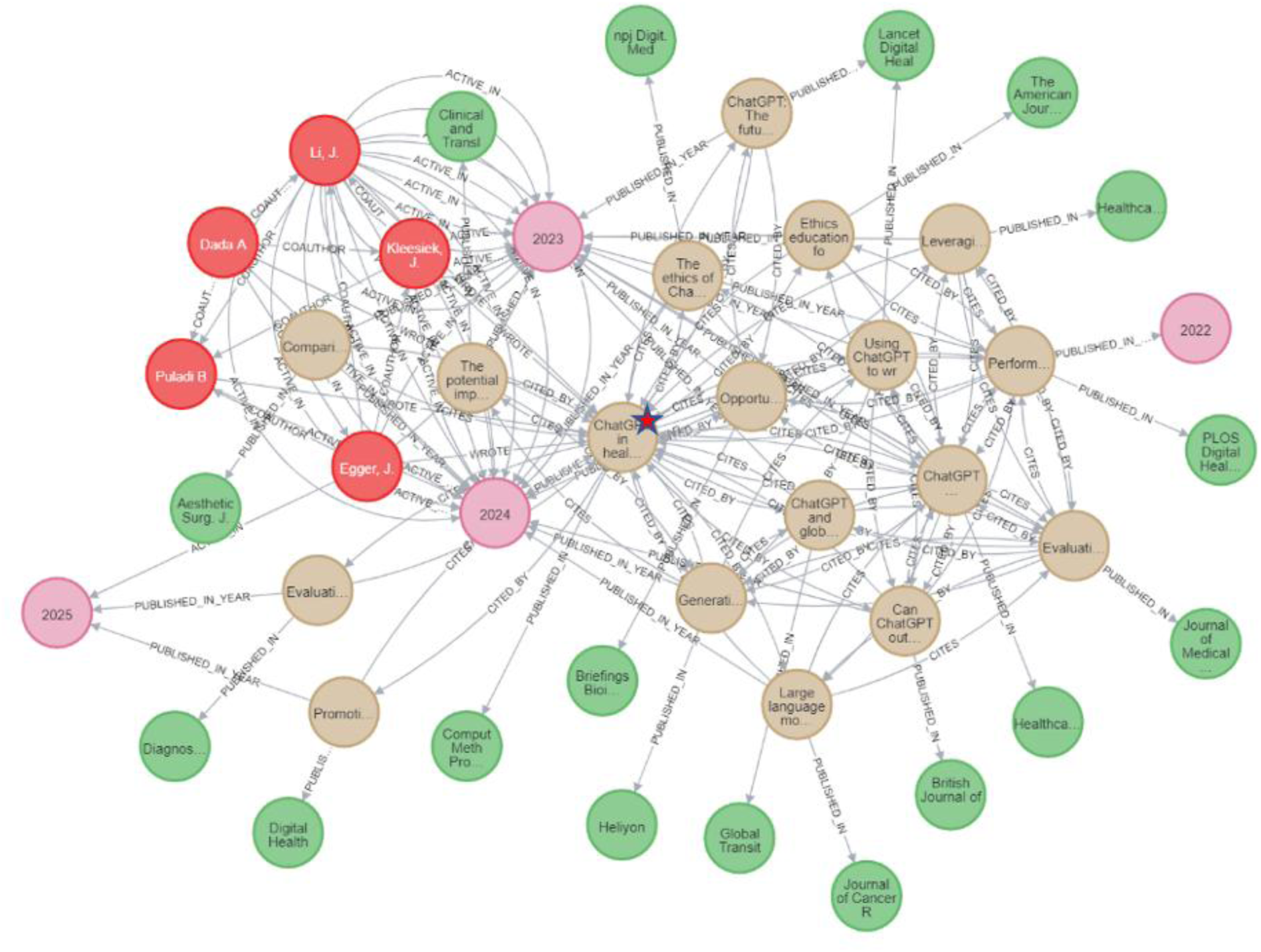
An illustrative example of a knowledge graph centered on a single focal article red node: Author; green node: Journal; pink node: Year; brown node: Article

## Discussion

### Behavior of LLM-based agentic AI

#### LLM agent vs. Human

In the study, an agentic AI workflow leveraged LLMs’ Natural Language Understanding (NLU) and Natural Language Generation (NLG) capabilities to identify metadata and explore trends in ChatGPT applications. NLU focuses on grasping the meaning of given text, whereas NLG involves automatically producing readable text from data using meaningful phrases and sentences [45]. However, compared with experts, who can provide more contextually relevant reviews, ChatGPT occasionally produces irrelevant or incorrect information [46]. Nevertheless, even when prompts were not grammatically correct, LLM agents captured semantic intent and provided appropriate responses, reflecting a form of human-like communication observed in the agentic AI workflow.

However, when communication breaks down, as it often does, LLM agents may display rigidity and make assertions without following through. From a probabilistic perspective, LLMs generate responses by sampling from high-probability regions of a conditional knowledge distribution. While the outputs tend to appear, in effect, like “average/mean” answers, which LLMs proffered. Prompt engineering and RAG constrain the probability distribution by introducing task-specific context with external knowledge, guiding the model toward more precise, context-aware outputs.

These observations suggest that prompt engineering, RAG technologies, and databases, along with agent interaction management such as answer checking and confirming, play an essential role throughout the development process. Furthermore, prompt intention analysis, used to reconfirm user intent, can clarify the intended purpose during collaboration within an Agentic AI workflow.

#### The Emerging Role of LLM Agents

Medicine is a humane endeavor in which language facilitates key interactions among clinicians, researchers, and patients; however, contemporary AI models for medical and healthcare applications have largely failed to leverage language fully [37]. This study examines LLM agents’ abilities in content recognition and semantic understanding across multiple documents, as well as their communication behaviors, to help researchers and healthcare providers gain deeper insights into their interactive behavior. Compared with traditional machine learning clustering and NLP topic modeling methods, LLMs offer more intuitive and interpretable grouping and classification, particularly when combined with prompt engineering and multi-agent refinement strategies.

These strategies are valuable because natural language is inherently ambiguous and difficult to define. Given their human-like communication patterns, the LLM agent may serve as an interaction layer alternative to traditional Human-Machine Interface (HMI) or User Interface (UI) for decision support systems. Additionally, AI-generated content may be used as a bridge between data-centric AI and knowledge-centric AI, through content generation, knowledge integration, and human-AI co-creation [47]. This ability suggests that LLM agents could receive and format data across disparate information systems, helping resolve the persistent information silos faced by healthcare providers.

### ChatGPT Applications Trend in Healthcare

#### ChatGPT Applications

This study revealed that Clinical decision support accounted for the largest share of ChatGPT applications, followed by Education and Medical research, consistent with the findings reported by Wu *et al.* [48]. The observed knowledge flow indicates a progressive shift in research directions. Early scholarship primarily focused on LLM performance [39] and prompt tuning [37], followed by a wave of systematic reviews that synthesized applications and technical limitations [36, 40, 41]. More recently, the discourse has shifted toward the ethical implications of real-world deployment, moving from technology-centric investigations to user- and society-focused considerations. Furthermore, as technologies advance, increasing emphasis has been placed on the safe and effective deployment of LLMs in clinical settings that require human expertise and where collaborative efforts outshine individual performance [30, 49, 50]. In this context, an adaptive, iterative refinement process of an agentic AI workflow that aligns with human oversight may contribute to more efficient, robust, and interpretable AI-enabled medical informatics studies.

Furthermore, studies show that applications are not confined to single-function use cases or domains but are increasingly expanding in scope and depth across diverse practices. At the same time, these applications are accompanied by a growing set of associated concerns, usage factors, and rapidly evolving applied techniques. Taken together, this indicates that focusing on any single dimension (such as ethics, safety, privacy, reliability, explainability, autonomy, or specific applied techniques) in isolation is insufficient. Additionally, this calls for an overarching governance approach that systematically addresses the integration of applied technological advancements, the rapid expansion of applications, and the associated concerns from a management AI perspective to manage AI challenges, risks, and technology effectively.

#### Associated concerns, factors influencing usage, and applied techniques

Over time, concerns about the data and knowledge limitations of LLMs have declined, suggesting these issues have become well recognized rather than focal points of ongoing analysis. In contrast, concerns related to human–AI collaboration and trust demonstrate a gradual increase, reflecting a shift in research emphasis from model-level limitations to interaction- and system-level challenges as LLMs are progressively integrated into real-world applications. The substantial overlap between the specific concern types and the top factors influencing usage indicates that issues, such as hallucination, trust, interpretability, transparency, bias, uncertainty, and ethics, simultaneously act as both drivers and constraints of LLM adoption.

From 2025 onward, factors not ranked among the top 10 began to appear, including autonomy, emergent behavior, and ambiguity. These developments align with the need for RAG and prompt engineering, alongside a growing focus on “Agent and Multi-agent” and “Agentic AI” techniques. Overall, the study suggests that ChatGPT application development is moving from emergence to a growth phase, characterized by the mutually reinforcing evolution of applied techniques, application domains, and factors influencing usage, while the maturity phase remains in its early stages.

#### Over-reliance

For individual use, LLM agents can serve as an auxiliary tool, helping with tasks such as text mining, knowledge understanding, and documentation, thereby saving time and increasing efficiency. However, as advances in AI technology enhance the generative capabilities of LLM agents and make errors harder to detect, users may become increasingly dependent on the answers they provide. Furthermore, in human–AI interaction, LLMs have been observed to tend to agree with users’ viewpoints, potentially reinforcing users’ existing beliefs and increasing the risk of overreliance. Although the study found that overreliance was reported in only 4 of 192 papers across 368 factors influencing usage, scholars have recently paid attention to the risks of overreliance and to preventing overdependence [51, 52]. Such overreliance may cause problems and could diminish critical thinking, leading users to lose their style and rely solely on LLMs. To mitigate unconscious overreliance, human or AI agents should know how to verify LLM answers that align with the user’s intended meaning and requirements. In addition, systematically recording generated responses in structured databases, such as a knowledge graph, may enable knowledge retrieval and traceability [53], facilitate inference and explanation [54, 55], visualize outputs [54, 55], and support reflective evaluation of human–AI collaboration.

### Limitations and Future Research

This study encountered several limitations during data extraction, identification, and synthesis. First, variable publication date labels and locations across articles were difficult to identify. As a result, in some cases, accepted dates were used instead of publication dates, suggesting that rule-based prompting may be needed. Second, existing PDF parsers still struggle to accurately interpret graphics and certain tables. However, these constraints do not undermine the study’s examination of agentic AI’s ability to identify, classify, and synthesize extracted data, nor the analysis of ChatGPT application trends by experts.

From a medical informatics perspective, LLMs demonstrate strong potential as text-mining tools for extracting information from unstructured academic literature; however, their probabilistic nature introduces non-deterministic synonyms. In addition, heterogeneous terminology and reference formats across the papers further complicate consistent data identification and integration. In this study, LLMs were used as normalization layers to harmonize citation structure and standardize terminology, thereby improving consistency in downstream analyses. To enhance reproducibility and semantic stability, future research should incorporate standard libraries (such as UMLS and ICD-9/10) and rule-based procedures that follow the LLM normalization layer to standardize terminology. Such hybrid approaches may support more deterministic vocabulary alignment, particularly for domain-specific factors that are not explicitly predefined.

While this study implemented a human–agentic AI collaboration workflow for metadata extraction, contextual synthesis, citation network construction, and database integration through iterative refinement, we focused primarily on output quality and result reproducibility for trend analysis. Formal evaluation of the agentic system was not conducted, which is a key limitation of the present work. The system-level performance and scalability of the agentic AI workflow were beyond the scope of this study and warrant further investigation.

The rapid expansion of LLMs and ChatGPT applications, along with the exponential growth of their user base, has far outpaced traditional IT systems. Moreover, unlike traditional IT systems, which yield consistent outputs, LLMs generate variable responses and produce hallucinations. These fundamental distinctions suggest that research on LLM adoption and sustained use may diverge significantly from conventional IT adoption frameworks. It is worthwhile to explore the factors driving their adoption and continued use to inform future practical implementation.

Using AI tools may increase efficiency and reduce the workload of healthcare professionals, allowing them to focus on higher-level tasks that require their expertise and clinical judgment [56]. Although integrating AI poses challenges, the potential collaboration between AI and physicians holds tremendous promise [49]. Lee *et al.* [9] suggest that these evolving tools can help healthcare providers deliver the best possible care, creating a more efficient, personalized, and patient-centric healthcare system [57]. Beyond being mere tools, LLM-based agents can dynamically adapt by adding tools as tasks require. We therefore suggest that agentic AI can be viewed as a decision-making assistant rather than merely a tool.

## Conclusion

We propose an approach that integrates an agentic AI workflow through literature review processes. The integration enables a broader range of NLP tasks, including extracting information from sparse data, reasoning and understanding, classification and clustering, and summarizing article content. Based on this method, the study provides a comprehensive overview of trends in ChatGPT applications, associated concerns, applied techniques, and usage factors in healthcare. The findings further provide meaningful insights into the operational mechanisms of agentic AI, demonstrating the potential of human–AI collaboration for medical informatics research.

## Ethical Approval

This study reviewed previously published studies, so no ethical approval or informed consent was necessary.

## Declaration of generative AI and AI-assisted technologies in the writing process

During the preparation of this manuscript, the authors used large language models (LLMs) to assist with data extraction and Grammarly to enhance language clarity. After using these tools, the authors reviewed and edited the content as needed and take full responsibility for the published article.

## CRediT authorship contribution statement

**Po Ju Chou:** Conceptualization, Data curation, Formal analysis, Investigation, Methodology, Software, Writing - original draft, Writing - review & editing.

**Yu Hsiu Lin:** Data curation, Formal analysis, Methodology, Validation, Visualization, Writing - review & editing.

**Eldon Y. Li:** Conceptualization, Formal analysis, Funding acquisition, Project administration, Supervision, Validation, Visualization, Writing - review & editing.

## Supplemental File(s)

A. List of ground-truth articles

B. Prompting strategies and classification categories for reference

C. Agentic AI Workflow Framework with a human-in-the-loop mechanism for reference

D. Extracted and identified metadata for reference

## Supporting information

List of ground-truth articles

Prompting strategies and classification categories for reference

Agentic AI Workflow Framework with a human-in-the-loop mechanism for reference

Extracted and identified metadata for reference

Extracted and identified metadata for reference

## Data Availability

All data produced in the present study are available upon reasonable request to the authors

## References

1. IBM Global AI Adoption Index 2022. Armonk, NY: IBM Corporation. May 2022. URL: https://www.ibm.com/downloads/cas/GVAGA3JP [accessed February 11 2026]

2. Loukides, M. AI Adoption in the Enterprise 2022. March 31, 2022. URL: https://www.oreilly.com/radar/ai-adoption-in-the-enterprise-2022/ [accessed February 11 2026]

3. Maslej N, Fattorini L, Perrault R, Gil, Y, Parli V, Kariuki N, et al. Artificial Intelligence Index Report 2025. arXiv preprint. 2035. [doi: 10.48550/arXiv.2504.07139]

4. Beam A. L, Drazen J. M, Kohane I. S, Leong T. Y, Manrai A. K, & Rubin E. J. Artificial Intelligence in Medicine. N Engl J Med. 2023;388(13):1220–1221. [doi: 10.1056/NEJMe2206291]

5. Ali S. R, Dobbs T. D, Hutchings H. A, & Whitaker I. S. Using ChatGPT to write patient clinic letters. Lancet Digit Health. 2023;5(4):e179–e181. [doi: 10.1016/S2589-7500(23)00048-1]

6. Cooper A, & Rodman A. AI and medical education: a 21st-century Pandora’s box. N Engl J Med. (2023);389(5):385–387. [doi: 10.1056/NEJMp2304993]

7. Liu J, Wang C, & Liu S. Utility of ChatGPT in clinical practice. J Med Internet Res. 2023; 25:e48568. [doi: 10.2196/48568]

8. Cascella M, Montomoli J, Bellini V, & Bignami E. Evaluating the Feasibility of ChatGPT in Healthcare: An Analysis of Multiple Clinical and Research Scenarios. J Med Syst. 2023;47(1):33. [doi: 10.1007/s10916-023-01925-4]

9. Lee P, Bubeck S, & Petro J. Benefits, limits, and risks of GPT-4 as an AI chatbot for medicine. N Engl J Med. 2023;388(13):1233–1239. [doi: 10.1056/NEJMsr2214184]

10. Liu S, Wright A. P, Patterson B. L, Wanderer J. P, Turer R. W, Nelson S. D, et al. Using AI-generated suggestions from ChatGPT to optimize clinical decision support. Journal of the American Medical Informatics Association. 2023;30(7):1237–1245. [doi: 10.1093/jamia/ocad072]

11. Rydzewski N. R, Dinakaran D, Zhao S. G, Ruppin E, Turkbey B, Citrin D. E, & Patel K. R. Comparative Evaluation of LLMs in Clinical Oncology. Nejm AI. 2024;1(5). [doi: 10.1056/AIoa2300151]

12. Sahni N. R, & Carrus B. Artificial Intelligence in U.S. Healthcare Delivery. N Engl J Med. 2023;389(4):348–358. [doi: 10.1056/NEJMra2204673]

13. Yao S, Zhao J, Yu D, Du N, Shafran I, Narasimhan K, & Cao Y. React: Synergizing reasoning and acting in language models. arXiv preprint. 2022. [doi: 10.48550/arXiv.2210.03629]

14. Hu Y, Chen Q, Du J, Peng X, Keloth V. K, Zuo X, et al. Improving large language models for clinical named entity recognition via prompt engineering. Journal of the American Medical Informatics Association. 2024;31(9):1812–1820. [doi: 10.1093/jamia/ocad259]

15. Nguyen J, & Pepping C. A. The application of ChatGPT in healthcare progress notes: A commentary from a clinical and research perspective. Clin Transl Med. 2023;13(7):e1324. [doi: 10.1002/ctm2.1324]

16. Zakka C, Shad R, Chaurasia A, Dalal A. R, Kim J. L, Moor, M, et al. Almanac - Retrieval-Augmented Language Models for Clinical Medicine. Nejm AI. 2024;1(2). [doi: 10.1056/aioa2300068]

17. Sapkota R, Roumeliotis K. I, & Karkee M. AI Agents vs. Agentic AI: A Conceptual taxonomy, applications and challenges. Information Fusion. 2025;126. [doi: 10.1016/j.inffus.2025.103599]

18. Gridach M, Nanavati J, Abidine K. Z. E, Mendes L, & Mack C. Agentic AI for scientific discovery: A survey of progress, challenges, and future directions. arXiv preprint. 2025. [doi: 10.48550/arXiv.2503.08979]

19. Acharya D. B, Kuppan K, & Divya B. Agentic AI: Autonomous Intelligence for Complex Goals—A Comprehensive Survey. IEEE Access. 2025;13:18912–18936. [doi: 10.1109/ACCESS.2025.3532853]

20. Xu F, Hao Q, Shao C, Zong Z, Li Y, Wang J, et al. Toward large reasoning models: A survey of reinforced reasoning with large language models. Patterns. (2025); 6(10):101370. [doi: 10.1016/j.patter.2025.101370]

21. Dayu Yang, Tianyang Liu, Daoan Zhang, Antoine Simoulin, Xiaoyi Liu, Yuwei Cao, Zhaopu Teng, Xin Qian, Grey Yang, Jiebo Luo, and Julian McAuley. Code to Think, Think to Code: A Survey on Code-Enhanced Reasoning and Reasoning-Driven Code Intelligence in LLMs. In Proceedings of the 2025 Conference on Empirical Methods in Natural Language Processing. 2025; 2586–2616, Suzhou, China. Association for Computational Linguistics. [doi: 10.18653/v1/2025.emnlp-main.130]

22. Pati A. K. Agentic AI: A Comprehensive Survey of Technologies, Applications, and Societal Implications. IEEE Access. 2025;13:151824–151837. [doi: 10.1109/access.2025.3585609]

23. Matsumoto N, Moran J, Choi H, Hernandez M. E, Venkatesan M, Wang P, & Moore J. H. KRAGEN: a knowledge graph-enhanced RAG framework for biomedical problem solving using large language models. Bioinformatics. 2024;40(6):btae353. [doi: 10.1093/bioinformatics/btae353]

24. Haug C. J, & Drazen J. M. Artificial Intelligence and Machine Learning in Clinical Medicine, 2023. N Engl J Med. 2023;388(13):1201–1208. [doi: 10.1056/NEJMra2302038]

25. Ferryman K, Mackintosh M, & Ghassemi M. Considering Biased Data as Informative Artifacts in AI-Assisted Healthcare. N Engl J Med. 2023;389(9):833–838. [doi: 10.1056/NEJMra2214964]

26. Hunter D. J, & Holmes, C. Where Medical Statistics Meets Artificial Intelligence. N Engl J Med. 2023;389(13):1211–1219. [doi: 10.1056/NEJMra2212850]

27. Dharanikota, Spurthy and Marakas, George M. Does AI Reliance Lead to Performance? A Task-Technology Fit Theory Perspective. AMCIS 2021 Proceedings. 21; 2021 August 9-13. [FREE full text]

28. He J, Baxter S. L, Xu J, Xu, J, Zhou X, & Zhang K. The practical implementation of artificial intelligence technologies in medicine. Nature Medicine. 2019;25(1):30–36.

29. Miao J, Thongprayoon C, Suppadungsuk S, Garcia Valencia O. A, & Cheungpasitporn W. Integrating retrieval-augmented generation with large language models in nephrology: advancing practical applications. Medicina. 2024;60(3):445. [doi: 10.3390/medicina60030445]

30. Pool J, Indulska M, & Sadiq S. Large language models and generative AI in telehealth: a responsible use lens. Journal of the American Medical Informatics Association. 2024;31(9):2125–2136. [doi: 10.1093/jamia/ocae035]

31. Lai H, Ge L, Sun M, Pan B, Huang J, Hou L, Yang Q, et al. Assessing the Risk of Bias in Randomized Clinical Trials With Large Language Models. JAMA Network Open. 2024;7(5):e2412687. [doi: 10.1001/jamanetworkopen.2024.12687]

32. Alharbi A, Hai A. A, Aljurbua R, & Obradovic Z. AI-Driven Sentiment Trend Analysis: Enhancing Topic Modeling Interpretation with ChatGPT. Artificial Intelligence Applications and Innovations. AIAI 2024; 2024:3–17. [doi: 10.1007/978-3-031-63215-0_1]

33. Carchiolo V, & Malgeri M. Trends, Challenges, and Applications of Large Language Models in Healthcare: A Bibliometric and Scoping Review. Future Internet. 2025;17(2). [doi: 10.3390/fi17020076]

34. Indrihapsari Y, Cahyadi Y, & Chung M.-Y. Emerging Trends in ChatGPT Research: A Quantitative Literature Review and Clustering Analysis. Journal of Advanced Research Design. 2025;142(1):51–77. [doi: 10.37934/ard.142.1.5177]

35. Olawade D. B, Plabon S. B, Ojo A, Ogunbona M. A, Makanjuola B. D, & Olasilola O. R. Human in the loop artificial intelligence in healthcare: applications, outcomes, and implementation challenges. Int J Med Inform. 2026;213:106362. [doi: 10.1016/j.ijmedinf.2026.106362]

36. Li J, Dada A, Puladi B, Kleesiek J, & Egger J. ChatGPT in healthcare: A taxonomy and systematic review. Comput Methods Programs Biomed. 2024;245:108013. [doi: 10.1016/j.cmpb.2024.108013]

37. Singhal K, Azizi S, Tu T, Mahdavi S. S, Wei J, Chung H. W, et al. Large language models encode clinical knowledge. Nature. 2023;620(7972):172–180. [doi: 10.1038/s41586-023-06291-2]

38. Jeblick K, Schachtner B, Dexl J, Mittermeier A, Stuber A. T, Topalis J, et al. ChatGPT makes medicine easy to swallow: an exploratory case study on simplified radiology reports. Eur Radiol. 2024;34(5):2817–2825. [doi: 10.1007/s00330-023-10213-1]

39. Kung T. H, Cheatham M, Medenilla A, Sillos C, De Leon L, Elepano C, et al. Performance of ChatGPT on USMLE: Potential for AI-assisted medical education using large language models. PLOS Digit Health. 2023;2(2):e0000198. [doi: 10.1371/journal.pdig.0000198]

40. Sallam M. ChatGPT Utility in Healthcare Education, Research, and Practice: Systematic Review on the Promising Perspectives and Valid Concerns. Healthcare (Basel). 2023;11(6):887. [doi: 10.3390/healthcare11060887]

41. Khan R. A, Jawaid M, Khan A. R, & Sajjad M. ChatGPT - Reshaping medical education and clinical management. Pak J Med Sci. 2023;39(2):605–607. [doi: 10.12669/pjms.39.2.7653]

42. Rahimzadeh V, Kostick-Quenet K, Blumenthal Barby J, & McGuire A. L. Ethics education for healthcare professionals in the era of ChatGPT and other large language models: Do we still need it? The American journal of bioethics. 2023;23(10):17–27. [doi: 10.1080/15265161.2023.2233358]

43. Ning Y, Teixayavong S, Shang Y, Savulescu J, Nagaraj V, Miao D, et al. Generative artificial intelligence and ethical considerations in healthcare: a scoping review and ethics checklist. Lancet Digit Health. 2024;6(11):e848–e856. [doi: 10.1016/S2589-7500(24)00143-2]

44. Sallam M, Al-Mahzoum K, Alaraji H, Albayati N, Alenzei S, AlFarhan F, et al. Apprehension toward generative artificial intelligence in healthcare: a multinational study among health sciences students. Frontiers in Education. 2025;10:1542769. [doi: 10.3389/feduc.2025.1542769]

45. Ali A. A. S, & Shandilya V. K. AI-Natural Language Processing (NLP). International Journal for Research in Applied Science and Engineering Technology. 2021;9(VIII):135–140. [doi: 10.22214/ijraset.2021.37293]

46. Mostafapour M, Fortier J. H, Pacheco K, Murray H, & Garber G. Evaluating Literature Reviews Conducted by Humans Versus ChatGPT: Comparative Study. JMIR AI. 2024;3:e56537. [doi: 10.2196/56537]

47. Ye J. Artificial intelligence-generated content (AIGC) in biomedical research, healthcare delivery, and clinical practices: technologies, applications, and regulatory considerations. Artificial Intelligence Review. 2026;59(2). [doi: 10.1007/s10462-025-11487-1]

48. Wu J, Ma Y, Wang J, & Xiao M. The Application of ChatGPT in Medicine: A Scoping Review and Bibliometric Analysis. J Multidiscip Healthc. 2024;17:1681–1692. [doi: 10.2147/JMDH.S463128]

49. Katz U, Cohen E, Shachar E, Somer J, Fink A, Morse E, Shreiber B, & Wolf, I. GPT versus Resident Physicians — A Benchmark Based on Official Board Scores. Nejm AI. 2024);1(5). [doi: 10.1056/AIdbp2300192]

50. Luo X, Tahabi F. M, Marc T, Haunert L. A, & Storey S. Zero-shot learning to extract assessment criteria and medical services from the preventive healthcare guidelines using large language models. Journal of the American Medical Informatics Association. 2024;31(8):1743–1753. [doi: 10.1093/jamia/ocae145]

51. Fatima A, Shafique M. A, Alam K, Fadlalla Ahmed T. K, & Mustafa M. S. ChatGPT in medicine: A cross-disciplinary systematic review of ChatGPT’s (artificial intelligence) role in research, clinical practice, education, and patient interaction. Medicine (Baltimore). 2024;103(32):e39250. [doi: 10.1097/MD.0000000000039250]

52. Xu T, Weng H, Liu F, Yang L, Luo Y, Ding Z, & Wang Q. Current Status of ChatGPT Use in Medical Education: Potentials, Challenges, and Strategies. J Med Internet Res. 2024;26:e57896. [doi: 10.2196/57896]

53. Yu T, Li J, Yu Q, Tian Y, Shun X, Xu L, Zhu L, & Gao H. Knowledge graph for TCM health preservation: Design, construction, and applications. Artif Intell Med. 2017; 77:48–52. [doi: 10.1016/j.artmed.2017.04.001]

54. Gao F, Zhao X, Xia D, Zhou Z, Yang R, Lu J, et al. Healthgenie: Empowering users with healthy dietary guidance through knowledge graph and large language models. arXiv preprint. 2025. [doi: 10.48550/arXiv.2504.14594]

55. Rajabi E, & Kafaie S. Knowledge Graphs and Explainable AI in Healthcare. Information. 2022;13(10). [doi: 10.3390/info13100459]

56. Xue V. W, Lei P, & Cho W. C. The potential impact of ChatGPT in clinical and translational medicine. Clinical and Translational Medicine. 2023;13(3):e1216. [doi: 10.1002/ctm2.1216]

57. Li J, Zhou Z, Lyu H, & Wang Z. Large language models-powered clinical decision support: enhancing or replacing human expertise? Intelligent Medicine. 2025;5(1):1–4. [doi: 10.1016/j.imed.2025.01.001]

