## Supplementary material for "Human–Agentic AI Collaboration Workflow for Trend Analysis of ChatGPT Applications in Healthcare": List of ground-truth articles

**Appendix A:** **List of ground-truth articles.**

a. 2023-H1: January 2023 ~ June 2023 (9 papers):

| Date | Author | Title | Description | Application | Journal |
| --- | --- | --- | --- | --- | --- |
| Feb.  06, 2023 | Patel & Lam | ChatGPT: the future of discharge summaries? | Discuss the usage and the future of ChatGPT in writing clinical discharge summaries. | Discharge summary | The Lancet Digital Health |
| Feb.  21, 2023 | Xue et al. | The potential impact of ChatGPT in clinical and translational medicine | Discuss the potential impact of ChatGPT in clinical and translational medicine. | Diagnosis and treatment of mental illness | Clinical and Translational Medicine |
| March 7, 2023 | Ali et al. | Using ChatGPT to write patient clinic letters. | Describe the early adoption and evaluation of ChatGPT-generated clinical letters to patients with limited clinical input. Aim to evaluate the readability, factual correctness, and humanness of ChatGPT-generated clinical letters to patients, using the example of skin cancer as the most common human cancer. | Patient clinic letters | The Lancet Digit Health |
| March 30, 2023 | Lee et al. | Benefits, limits, and risks of GPT-4 as an AI chatbot for medicine | Describe another type of AI, the medical AI chatbot. Benefits, limits, and risks of GPT-4 as an AI chatbot for medicine. | Medical note-taking,  Innate medical knowledge,  Medical consultation, | New England Journal of Medicine (NEJM) |
| March 30, 2023 | Haug et al. | Artificial Intelligence and Machine Learning in Clinical Medicine, 2023 | Introducing the progress in data science, AI, and machine learning in medicine, discussing unresolved issues in these areas, and exploring the issues of chatbots in medicine. | Medical documentation,  Clinical note | NEJM |
| April 22, 2023 | Liu, S.,  et al. | Using AI-generated suggestions from ChatGPT to optimize clinical decision support | To determine if ChatGPT can generate useful suggestions for improving clinical decision support (CDS) logic and to assess noninferiority compared to human-generated suggestions. | Clinical alert suggestions | Journal of the American Medical Informatics Association (JAMIA) |
| Jun 15,  2023 | Rodman et al. | Accuracy of a Generative Artificial Intelligence Model in a Complex Diagnostic Challenge | This study assessed the performance of Generative Pre-trained Transformer 4 (GPT-4) in clinical applications, such as complex diagnostic reasoning, using the New England Journal of Medicine clinical-pathologic conferences. | Complex diagnostic reasoning | JAMA Network Open |
| June 25, 2023 | Nguyen &  Pepping | The application of ChatGPT in healthcare progress notes: A commentary from a clinical and research perspective | Commentary article using clinical and research perspectives on the application of ChatGPT in healthcare progress notes.  Outline the potential benefits and pitfalls of implementing ChatGPT to create and manage healthcare progress notes using prompt engineering techniques. | Progress notes | Clinical and Translational Medicine |
| June 28, 2023 | Liu, J.,  et al. | Utility of ChatGPT in clinical practice | To better understand the application of ChatGPT in clinical practice, this study reviewed recent advances to help researchers effectively grasp key aspects of the topic and identify possible future research directions.  ChatGPT is utilized in clinical practice to support decision-making, provide reliable information for medical question-answering, and aid documentation.  Future research explores real-time monitoring, predictive analytics, precision medicine, telemedicine, and integration with healthcare systems. | Clinical decision support,  Medical question-answering,  Medical documentation | Journal of Medical Internet Research |

b. 2023-H2: July 2023 ~ December 2023 (6 papers)

| Date | Author | Title | Description | Application | Journal |
| --- | --- | --- | --- | --- | --- |
| July 12, 2023 | Singhal et al. | Large language models encode clinical knowledge | PaLM and its instruction-tuned variant, Flan-PaLM, have achieved state-of-the-art accuracy across various medical question-answering datasets. However, despite improvements in comprehension, knowledge recall, and reasoning with increased model scale and instruction prompt tuning, human evaluations reveal that these models still fall short of clinician performance, highlighting the need for further research to develop safe and effective LLMs for clinical use. | Multiple-choice questions,  Medical question answering | Nature |
| July 27, 2023  (Aug 3  , 2023) | Sahni & Carrus | Artificial Intelligence in U.S. Health Care Delivery | Examining the use of AI in reimbursement, clinical operations, quality, and safety, and discussing the challenges (22 factors in 6 categories) that healthcare organizations face in deploying AI. | Insurance reimbursement, clinical operations, quality, and safety | NEJM |
| Nov. 6,  2023 | Li & Zheng | Identification of Ancient Chinese Medical Prescriptions and Case Data Analysis Under Artificial Intelligence GPT Algorithm: A Case Study of Song Dynasty Medical Literature. | Combined with AI prescription recognition and ChatGPT technology, medical data mining was conducted to obtain information and knowledge on medical research in the Song Dynasty, understand the clinical practice experience of ancient Chinese medicine, and to provide a historical reference for the clinical application and research of modern Chinese medicine. | Ancient Chinese medicine prescriptions,  Medical named entity recognition (NER) | IEEE Access |
| Nov. 9,  2023 | Eriksen et al. | Use of GPT-4 to Diagnose Complex Clinical Cases | Assessed the performance of the newly released AI GPT-4 in diagnosing complex medical case challenges and compared the success rate to that of medical journal readers.  GPT-4 correctly diagnosed 57% of cases, outperforming 99.98% of simulated human readers generated from online answers | Complex real-life medical cases | NEJM AI |
| Nov. 17, 2023 | Benary et al. | Leveraging Large Language Models for Decision Support in Personalized Oncology | To assess performance and define their role using four recent LLMs as support tools for precision oncology.  This diagnostic study examined 10 fictional cases of  patients with advanced cancer with genetic alterations. Each case was submitted to 4 LLMs and one expert physician to identify personalized treatment options. | Decision support | JAMA Network Open |
| Dec. 26,  2023 | Hu et al. | Improving large language models for clinical named entity recognition via prompt engineering | The study highlights the potential of large language models, specifically GPT-3.5 and GPT-4, to process complex clinical data and extract meaningful information with minimal training data. By developing and refining prompt-based strategies, we can significantly enhance the models' performance, making them viable tools for clinical NER tasks and possibly reducing the reliance on extensive annotated datasets. | Clinical named entity recognition (NER),  Clinical notes | JAMIA |

c. 2024-H1: January 2024 ~ June 2024 (12 papers)

| Date | Author | Title | Description | Application | Journal |
| --- | --- | --- | --- | --- | --- |
| Jan. 17,  2024 | Wu et al. | Benchmarking Open-Source Large Language Models, GPT-4 and Claude 2 on Multiple-Choice Questions in Nephrology | This study investigated the medical knowledge capability of multiple LLMs in nephrology by evaluating their performance on nephSAP multiple-choice questions. Open-source LLMs achieved success rates of 17.1% to 30.6%, whereas Claude 2 and GPT-4 achieved 54.4% and 73.3%, respectively, while the passing grade for human test takers was 75%. | Multiple-Choice Questions in Nephrology | NEJM AI |
| Jan. 25,  2024 | Zakka et al. | Almanac - Retrieval-Augmented Language Models for Clinical Medicine | LLMs' adoption has been limited by their tendency to generate incorrect and sometimes even harmful statements. To address this issue, this study tasked a panel of 8 board-certified clinicians and two healthcare practitioners with evaluating Almanac, an LLM framework augmented with retrieval capabilities from curated medical resources for medical guidelines and treatment recommendations. The panel compared responses from Almanac and standard LLMs against a novel dataset of 314 clinical questions spanning nine medical specialties.  Almanac significantly improved performance compared with the standard LLMs across factuality, completeness, user preference, and adversarial safety axes. | Clinical questions answering | NEJM AI |
| Feb. 14, 2024 | Pool  & Sadiq | Large language models and generative AI in telehealth: a responsible use lens | To explore "What is the current state of research on the responsible use of LLMs in telehealth, and what are the knowledge gaps in this context of use?", this study conducted a scoping review following the methodological framework established by Arksey and O'Malley.  This review assesses the current research landscape of the application and use of LLMs and generative AI through ChatGPT in telehealth. Additionally, the review seeks to identify key areas for future research, focusing on AI ethics considerations for responsible use and ensuring trustworthy AI. | Telehealth | JAMIA |
| March 8, 2024 | Miao et al. | Integrating retrieval-augmented generation with large language models in nephrology: advancing practical applications | This review explores the potential application of LLMs integrated with RAG in nephrology and analyzes its strengths and weaknesses. | Chronic kidney disease | Medicina |
| April 12, 2024 | Katz et al. | GPT versus Resident Physicians — A Benchmark Based on Official Board Scores | This study evaluated the performance of GPT-3.5 and GPT-4 on the 2022 Israeli board residency examinations and compared the results with those of 849 practicing physicians.  GPT-4 ranked higher than the majority of physicians in psychiatry, with a median percentile of 74.7%, and it performed similarly to the median physician in general surgery and internal medicine, displaying median percentiles of 44.4% and 56.6%, respectively.  GPT-4 performance was lower in pediatrics and OB/GYN but remained higher than a considerable fraction of practicing physicians, with median scores of 17.4% and 23.44%, respectively. | Israeli board residency examinations,  Multiple-choice questions (a, b, c, or d), | NEJM AI |
| April 16, 2024 | Rydzewski  et al. | Comparative Evaluation of LLMs in Clinical Oncology | This study tested five publicly available LLMs on a comprehensive battery of 2044 oncology questions.  There was significant heterogeneity in performance between models (P<0.001). Relative to a human benchmark (2013 and 2014 examination results), GPT-4 was the only model to perform above the 50th percentile. | oncology questions | NEJM AI |
| May. 08, 2024 | Skryd & Lawrence | ChatGPT as a Tool for Medical Education and Clinical Decision-Making on the Wards: Case Study | This study explores ChatGPT as a real-time interactive educational tool during inpatient teaching rounds at a large urban academic medical center. It identifies key use cases and qualitatively assesses their impact on residents' and attendings' clinical learning, diagnostic reasoning, and medical decision-making. | Medical education,  Clinical decision-making | JMIR Formative Research |
| May 22, 2024 | Lai et al. | Assessing the Risk of Bias in Randomized Clinical Trials With Large Language Models | In this survey study of applying LLMs for ROB assessment, LLMs demonstrated substantial accuracy and consistency in evaluating RCTs, suggesting their potential as supportive tools in systematic review processes. | ROB in randomized clinical trials (RCTs) | JAMA Network Open |
| May 30, 2024 | Liu et al. | Using large language model to guide patients to create efficient and comprehensive clinical care message | This study investigated the feasibility of using LLMs to engage with patients at the time they are drafting a question to their healthcare providers and generate pertinent follow-up questions that the patient can answer before sending their message to ensure that their healthcare provider receives all the information they need to safely and accurately answer the patient's question, eliminating back-and-forth messaging, and the associated delays and frustrations.  The fine-tuned CLAIR model had the best performance. The GPT4 model received higher scores for utility and completeness but lower scores for clarity and conciseness. CLAIR generated follow-up questions with similar clarity and conciseness as the actual follow-ups written by healthcare providers, with higher utility than healthcare providers and GPT4, and lower completeness than GPT4, but better than healthcare providers. | follow-up questions | JAMIA |
| June 3,  2024 | Luo et al. | Zero-shot learning to extract assessment criteria and medical services from the preventive healthcare guidelines using large language models | This study investigates the feasibility of using LLMs to automate the extraction of assessment criteria and risk factors from the guidelines for future analysis of medical records in EHRs.  24 guidelines were included in this study. The LLMs can automate the extraction of all criteria, risk factors, and medical services from 9 guidelines. All 3 LLMs perform well in extracting demographic criteria or risk factors information. Some LLMs perform better on extracting the social determinants of health, family history, and preventive counseling services than others. | criteria and risk factors in the USPSTF guidelines,  clinical NER, | JAMIA |
| June 17, 2024 | Unlu et al. | Retrieval-Augmented Generation–Enabled GPT-4 for Clinical Trial Screening | Traditionally, participant screening in clinical trials is a labor-intensive, error-prone process that requires significant time and resources, so leveraging LLMs such as GPT-4 offers an opportunity to enhance screening.  This study evaluated the utility of an RAG–enabled GPT–4 to improve the accuracy, efficiency, and reliability of screening for a trial involving patients with symptomatic heart failure. The sensitivity and specificity for determining patient eligibility with Inclusion Exclusion Review were 92.3% and 93.9%, respectively, and 90.1% and 83.6% with the study staff. | patient clinical trial screening | NEJM AI |
| Jun. 20, 2024 | Lo et al. | Dietary Assessment With Multi-modal ChatGPT: A Systematic Analysis | This study uses food images and guidance prompts as inputs to multi-modal ChatGPT (GPT-4V), enabling it to identify food items, estimate their weight, determine their source, and provide dietary recommendations. | Dietary assessment | IEEE Journal of Biomedical and Health Informatics |

d. 2024-H2: July 2024 ~ December 2024 (8 papers)

| Date | Author | Title | Description | Application | Journal |
| --- | --- | --- | --- | --- | --- |
| July. 19,  2024 | Fatima et al. | ChatGPT in medicine: A cross-disciplinary systematic review of ChatGPT's (artificial intelligence) role in research, clinical practice, education, and  patient interaction | Following the PRISMA guidelines, a systematic search of PubMed, Medline, Embase, and Scopus identified 83 studies for inclusion in this review, which provides a comprehensive overview of ChatGPT's capabilities and limitations in medical science, healthcare research and education, scientific writing, and clinical practice. | Healthcare education, research, writing, patient communication,  Practice | Medicine |
| Aug. 28,  2024 | Xu et al. | Current Status of ChatGPT Use in Medical Education: Potentials, Challenges, and Strategies | This study examines the potential and limitations of integrating ChatGPT into medical education by assessing its capabilities and challenges.  A systematic search of the Web of Science (January 2022–December 2024) was conducted. The extracted literature was then analyzed using CiteSpace v6.1R6 Basic to explore keyword trends and research patterns within the selected studies. | Medical Education | Journal of Medical Internet Research |
| Aug. 29,  2024 | Dergaa et al. | A Thorough Examination of ChatGPT-3.5 potential applications in medical writing: A preliminary study | This study evaluates ChatGPT's effectiveness in medical writing, focusing on its potential to enhance clarity, efficiency, and reliability in presenting medical information. Through a thorough literature review, we identified the most relevant and impactful themes and tasks that ChatGPT can perform in medical writing. | Medical Writing | Medicine |
| Oct. 03, 2024 | Goktas & Grzybowski | Assessing the Impact of ChatGPT in Dermatology: A Comprehensive Rapid Review | This study analyzes ChatGPT's role in dermatology, specifically exploring its applications in clinical diagnostics, patient education, and medical writing while addressing the ethical and legal challenges associated with its use. A comprehensive search was conducted across Web of Science, Scopus, PubMed, and MEDLINE to identify relevant studies, focusing on articles published between November 2022 and August 2024. | Patient education, clinical decision support, medical writing | Journal of Clinical Medicine |
| Oct. 07, 2024 | Blanchard et al. | Generative AI-based knowledge graphs for the illustration and development of mHealth self-management content | This study aims to employ generative artificial intelligence (AI)-based knowledge graphs and network analysis to categorize and structure mHealth content at the example of a fibromyalgia self-management program. | Mobile health  Digital therapeutics (DTx) | Frontiers in Digital Health |
| Oct. 27, 2024 | Ghanta et al. | Applications of ChatGPT in Heart Failure Prevention, Diagnosis, Management, and Research: A Narrative Review | This study examined existing research on ChatGPT's use in heart failure (HF) care by searching PubMed, Google Scholar, Medline, and Scopus. It assessed ChatGPT's role in HF prevention, diagnosis, and management. The findings suggest that ChatGPT facilitates knowledge dissemination by providing accessible explanations of complex topics, potentially enhancing patient-clinician communication and bridging the knowledge gap. | Heart failure prevention, diagnosis,  Management, Research | Diagnostics |
| Nov 15, 2024 | Zhang et al. | Examining the Role of Large Language Models in Orthopedics: Systematic Review | This review comprehensively summarizes research findings on applying LLMs in orthopedics, highlighting their advantages, limitations, and methodological evaluations. It also explores emerging opportunities and challenges in this field, emphasizing their role in facilitating interdisciplinary collaboration and advancing research between computer science and orthopedics. | Clinical practice,  Education, Scientific research, management | Journal of Medical Internet Research |
| Dec. 09,  2024 | Zarfati et al. | Exploring the Role of Large Language Models in Melanoma:  A Systematic Review | This systematic review evaluates the current applications, advantages, and challenges of large language models (LLMs) in melanoma care. In accordance with PRISMA guidelines, the review was conducted using the PubMed and Scopus databases, focusing on LLM applications in melanoma. Nine studies were categorized into subgroups: patient education, diagnosis, and clinical management. LLMs demonstrated high accuracy in inpatient education, although readability often exceeded recommended levels. For diagnosis, multi-modal LLMs such as GPT-4V showed the ability to distinguish melanoma from benign lesions, but accuracy varied, influenced by factors such as image quality and the integration of clinical context. | Patient education, Diagnosis, Clinical management | Journal of Clinical Medicine |
