## Supplementary material for "Human–Agentic AI Collaboration Workflow for Trend Analysis of ChatGPT Applications in Healthcare": Prompting strategies and classification categories for reference

**Appendix B: Prompting strategies and classification categories**

1. **ChatGPT applications**

**prompt** = f"""

*You are an expert model specialized in medical text classification and summarization, capable of applying professional healthcare terminology, identifying key clinical concepts, and generating concise and academically structured outputs.*

*Analyze the following short descriptions of ChatGPT / LLM applications in healthcare:*

*{applications_text}*

*Classify each entry into one of the following categories (use the best fit):*

*1*. Clinical Decision Support*,*

*such as: Diagnostic support; Diagnosis prediction; Early Diagnosis of ophthalmologic diseases; Diagnosis and medical Q&A; Prescription entity recognition; Thyroid Disease Management; Treatment; Risk Assessments in mental health; Triage and Symptom Assessment; Remote Monitoring and Support; patient monitoring; remote patient monitoring; surgical oncology*

*2.* Medical Research and Data Analysis,

*such as: Assistance in research; Biomedical Research; Clinical data analysis; clinical trials; Drug Discovery; Medical research analysis; Scientific research; case study analysis; health care task analysis; improved data collection and analysis;*

*3.* Medical Documentation,

*such as: Clinical language understanding; Medical text summarization; Text clasification; Medical writing, Natural language interpretation; Language Translation; Medical notes; Clinical reports; Patient clinic letters; Progress notes; Discharge summaries; extracting structured data from clinical trial documents; information extraction; medical advice generation;*

*4*. Education (Medical and Health),

*such as: Health Education; Health Informatics; Medical Education; Physician Training; perioperative patient education;*

*5.* Healthcare/Medical Workflow,

*such as: Electronic Health Record (EHR)/EMR management; Data interoperability; Clinical trial recruitment; clinical data management; clinical management support; clinical practice; patient data management; patient outcomes improvement; patient care; direct patient care; Medical Copilot; detecting and communicating uncertainty; safety; patient management and treatment; ethical considerations in healthcare;*

*6.* Healthcare/Medical Consultation,

*such as: Patient communication; Patient interaction and assistants; Question answering; Medical question answering; Healthcare; Healthcare management; Medical information; Mental Healthcare Assistance; chronic disease management;*

*7.* Public Health

*For each text:*

*- Return the best-matching category.*

*- Provide a one-sentence rationale.*

*Also return a summarized list of unique major categories identified.*

*Return the entire result STRICTLY as JSON matching this schema:*

*{{*

*"classified_reasons": [*

*{{"text": "<original text>", "category": ["Category"], "rationale": "<one-sentence rationale>"}}*

*],*

*"application_categories": ["CategoryA", "CategoryB"]*

*}}*

*Do not output any additional explanation or commentary. Only output valid JSON.*

"""

2. **Applied techniques**

prompt = f"""

*You are an expert model specialized in Medical informatics and text classification and summarization.*

*Analyze the following short descriptions of ChatGPT / LLM applications in healthcare:*

*Text: {tech_text}*

*Rules must follow:*

*- Classify ONLY the technologies directly related to improving or enhancing the performance and adaptability of LLM-based systems.*

*- ⚠️ Exclude any unrelated or generic technologies such as:*

*"machine learning", "deep learning", "natural language processing (NLP)", "data mining", "AI in general", or other broad or traditional techniques not specifically referring to LLMs or their enhancement.*

*- If the text merely describes interactions between two sessions or instances of GPT without explicitly using an agent framework, DO NOT classify it as "Agent" or "Multi-Agent" category.*

*- DO NOT classify model name, such as "Generative Pre-trained Transformer 4 (GPT-4)" into Fine-Tuning category.*

*- If the text only focus on discusses/describe such as: "should explore fine-tuning", "LLMs often require fine-tuning to adapt to specific contexts and improve their performance", "suggest adjustments to model parameters or retraining to improve accuracy"), DO NOT classify into Fine-Tuning category.*

*Each technology MUST belong to one or more of the following five categories and definitions (choose only from these).*

*Categories and definitions:*

*1. **Prompt Engineering** – including few-shot prompting, chain-of-thought reasoning, and instruction refinement to optimize query formulation and task-specific responses. Iterative reasoning, standardized chat prompt, self-consistency checks, and corrective loops are applied to improve accuracy and reliability, closely related to prompt design.*

*2. **Retrieval-Augmented Generation** – RAG integrates external knowledge retrieval with LLM generation to enhance accuract/correctness and contextual relevance (rag, embedding search, information retrieval), such as: 'Data extraction and integration with existing health care systems' suggest the use of external knowledge to enhance LLM outputs.*

*3. **Agent and Multi-Agent** – using single or multiple agents to execute tasks in parallel or sequentially, via role specialization and tool integration, with collaborative interactions to achieve task goals. This design does not necessarily constitute a complete workflow.*

*4. **Agentic AI** – employing autonomous or collaborative agents (e.g., Data Agent, Code Agent, Evaluator Agent, Validator Agent) to facilitate task planning, agent utilization, iterative evaluation and validation of agents (iterative reasoning, self-consistency checks, and corrective loops with Agents) , and dynamic workflow orchestration to improve agent's accuracy and reliability.*

*5. **Fine-Tuning** – exclusively adjusting LLMs' parameters or retraining the language model using domain-specific data and terminology to align outputs with target contexts.*

*For each text:*

*- Provide a one or two sentences rationale.*

*- Also return a summarized list of unique major categories identified.*

*Return STRICTLY as JSON:*

*{{*

*"reason_techs": [*

*{{"category": ["CategoryA", "CategoryB"], "rationale": "<one-sentence rationale>"}}*

*],*

*"tech_categories": ["CategoryA", "CategoryA"]*

*}}*

*Do not output any additional explanation or commentary. Only output valid JSON.*

*Do not over explain/interpretation, such as:*

*- "prompting the system to validate its own outputs" is NOT necessary involves the use of autonomous agents;*

*- mention of 'large language models' NOT implies 'the use of agents to execute tasks';*

*- 'Pedagogical methodologies', 'AI tools' NOT imply 'the use of multiple agents to achieve specific tasks';*

*- 'error validation mechanisms', 'Human evaluation framework', 'transparency', 'mechanisms to address hallucinations and errors' NOT suggests 'the use of iterative evaluation and validation processes, which aligns with the concept of Agentic AI'.*

*- 'self-appraised confidence scores', 'task management tools', 'the facilitation of task planning and iterative evaluation' NOT indicates the use of autonomous or collaborative agents.*

*- 'mention of 'Cohen's kappa' relates to evaluating model performance' NOT involve fine-tuning,*

*- 'BioMedLM indicates a specialized model', Specialized ChatGPT model' NOT implies to fine-tuning.*

*fine-tuning exclusively adjusting LLMs' parameters or retraining the language model using domain-specif data.*

*Fine-tuning via prompt design, prompting techniques, RAG, concept matrix, model selection or other technologies/methods are not included in this category.*

"""

3. **Associated concerns**

#Extract Concerns (text span)

prompt = f"""

You are an expert in biomedical NLP, GenAI, and healthcare/medical informatics.

Extract terms, sentences or text spans that describe concerns, limitations, risks, barriers, or challenges related to ChatGPT applications in healthcare.

These include, but are not limited to: accuracy or probable inaccuracy, ethical issues, lack of emotional sensitivity, need for up-to-date data or limited data availability, concerns of plagiarism or misconduct, uncertainty, privacy or privacy protection, fairness or inclusiveness, communication or trust issues, security, data interpretation, replacing or replace of humans, inconsistency, accountability, governance, plausibility, hallucination or hallucinatory outputs, overreliance or overdependence, reliability, lack of transparency, or concerns regarding beneficial outcomes.

Rules:

- Extract the complete sentence or closest complete clause.

- Do NOT paraphrase.

- Do NOT classify.

- Do NOT summarize.

- Return raw text spans.

- If nothing relevant, return an empty list.

Return STRICTLY in this JSON schema:

{{

"extracted_concerns": [

"<text span 1>",

"<text span 2>"

]

}}

Text:

{text}

"""

#sub_category Classification

prompt = f"""

*Using medical and IT terminology, identify, extract, and describe the concerns in the extracted concerns text.*

*Then, assign appropriate sub_categories to each extracted concern,*

*ensuring that the sub_categories are expressed using healthcare-specific terminology (e.g., "data protection", "responsibility", "trust").*

*Output strictly in this JSON format:*

*{{"sub_categories": ["sub_categoryA", "sub_categoryB"]}}*

*Do NOT invent or infer.*

*Do NOT use generic terms or phrases (e.g., "limitations", "users solely responsible for errors") as a sub_category.*

*Extracted Concerns text:*

*{concerns_list}*

"""

#assign key six categories

**CATEGORY_SYSTEM_MESSAGE** = SystemMessage(*content="""*

*You are a strict classifier for concerns related to ChatGPT applications in healthcare.*

*Classify each sub_category into exactly ONE of the following six categories,*

*based on its PRIMARY concern.*

***Ethical, Safety, and Legal concerns**:*

*Assign if the concern primarily involves:*

*- ethical or moral issues (socoal bias, fairness, inclusiveness, autonomy)*

*- bias, safety risks or harm*

*- privacy, data leakage, data protection, confidentiality, or security*

*- legal responsibility, liability, governance, guideline adherence, or accountability*

*- plagiarism, misconduct, or copying*

*- overreliance on LLMs*

*- replacing human*

*Exclude if the issue is mainly about model accuracy, explainability, data quality,*

*or deployment logistics without explicit ethical, safety, or legal implications.*

***Reliability, Explainability and Interpretability challenges**:*

*Assign if the concern primarily involves:*

*- hallucination or fabricated content*

*- mixed-accuracy or incorrect responses*

*- inconsistency or instability*

*- lack of transparency or black-box behavior*

*- limited traceability, explainability, or interpretability*

*- uncertainty or implausibility of results*

*Exclude if the concern is about insufficient data, evaluation benchmarks, or ethical misuse.*

***Data and Knowledge limitations**:*

*Assign if the concern primarily involves:*

*- outdated medical or scientific knowledge*

*- limited, biased, or incomplete datasets*

*- insufficient domain-specific or local clinical data*

*- knowledge gaps due to training data constraints*

*Exclude if the focus is on validation, research gaps, therapeutic value, interpretation, guideline adherence, or ethical governance.*

***Evaluation and Validation deficiencies**:*

*Assign if the concern primarily involves:*

*- lack of standardized evaluation or validation methods, verification, performance evaluation*

*- absence of unified clinical benchmarks or gold standards*

*- difficulty validating multifactorial clinical outcomes*

*- inadequate external, prospective, or real-world validation*

*Exclude if the issue is mainly about reliability, data availability, or implementation feasibility.*

***Implementation and Knowledge-management barriers**:*

*Assign if the concern primarily involves:*

*- generalizability, research gaps, deployment or integration difficulties*

*- high infrastructure or maintenance costs*

*- low observed benefit or return on investment*

*- quality, poor user acceptance, training, or workflow alignment*

*- therapeutic value, inadequate knowledge-management practices*

*Exclude if the concern is about ethics, trust, performance evaluation, or model correctness.*

***Collaboration and Trust**:*

*Assign if the concern primarily involves:*

*- trust or distrust in AI-generated outputs*

*- trustworthy AI from the user or clinician perspective*

*- undermined communication*

*- insufficient human-in-the-loop or human review mechanisms*

*- weak collaboration or coordination frameworks*

*Exclude if the concern is fundamentally about ethical/legal accountability,technical reliability, or deployment constraints.*

*Rules:*

*- Do NOT output code fences.*

""")

#key Category Classification

prompt = f"""

*Classify each extracted sub_categories from Sub_Category into one category:*

*MANDATORY rules:*

*- Do NOT paraphrase the original text.*

*- Treat each sub_category as a separate item.*

*- Assign exactly one category per sub_category.*

*- when multiple sub_category values exist, treat each sub_category as a separate item,*

*and assign one corresponding category per sub_category, preserving the original order and count.*

*- If multiple sub_category values belong to the same category, **repeat that Category for each sub_category**,*

*instead of merging them.*

*- Provide one-sentence rationale per sub_category.*

*Return STRICTLY in this JSON schema.*

*Do NOT include comments, trailing commas, ```json, or any extra text:*

*{{*

*"concern_reasons": [*

*{{*

*"text": "<original text from Extracted Concerns text>",*

*"sub_category": ["sub_CategoryA", "sub_CategoryB"],*

*"category": ["corresponding CategoryA", "corresponding CategoryB"],*

*"rationale": ["<rationale for sub_CategoryA>", "<rationale for sub_CategoryB>"]*

*}}*

*]*

*}}*

*Extracted Concerns text:*

*{concerns_list}*

*Sub_Category:*

*{sub_category}*

"""

**Note:**

1. **Due to technical considerations, the content of the relevant prompts above has not been grammatically corrected.**
2. **Another purpose is to examine and refine the interactions and adaptability of LLM agents under unmodified human-like everyday conversation conditions.**
