## Supplementary material for "Human–Agentic AI Collaboration Workflow for Trend Analysis of ChatGPT Applications in Healthcare": Agentic AI Workflow Framework with a human-in-the-loop mechanism for reference

**Appendix C: Agentic AI Workflow Framework with a human-in-the-loop mechanism**

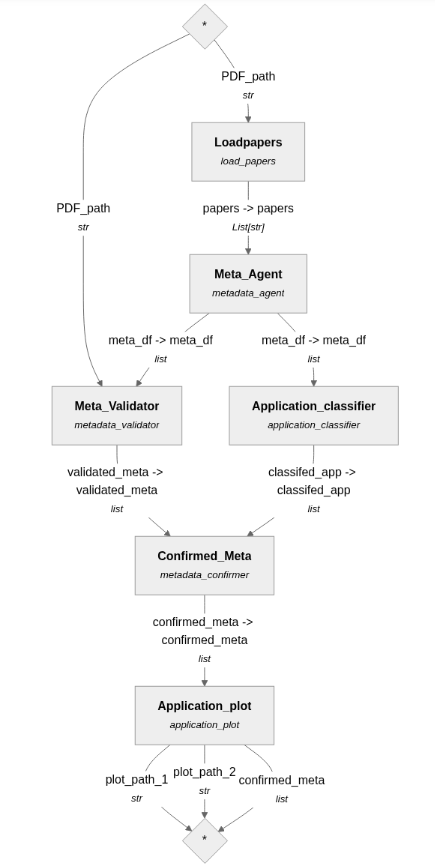
a.

**Function of Components**

***Loadpapers****:*

load PDF documents

***Meta_Agent****:*

LLM agent in charge of embedding loaded documents and extracting metadata

***Meta_Validator****:*

LLM judger, validating extracted metadata

***Application_classifier*:**

LLM agent classifying / clustering applications from documents

***Confirmed_Meta*:**

correct, and confirm metadata by LLM agent

***Application_Plot*:**

visualizing results of identified and classified applications, and saving images

**Note:** Each component can be examined separately, added to, and deleted as necessary by the human to meet the intended goal.

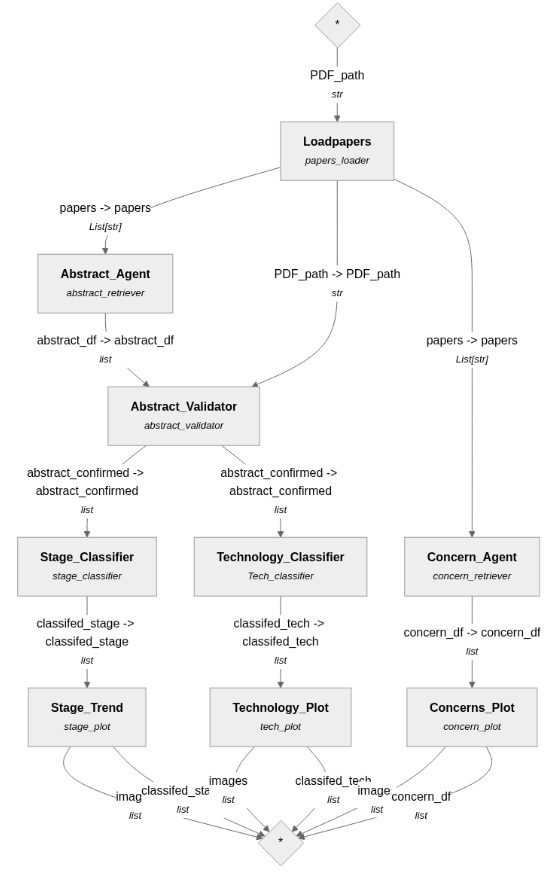
b.

***Legend:***

*Abstract_Agent:* response for abstract extraction

*Abstract_Validator:* validates and corrects the extracted abstract

*Stage_ Classifier:* classifying the development stage according to the extracted abstract

*Technology_Classifier:* classifies the applied techniques

*Concern_Agent:* includes 3 agents to extract associated concerns from the full text, and identify/confirm the results

*Stage_Trend, Technology_Plot, Concerns_Plot:* visualizing the trend of development stages, identified applied techniques, and identified associated concerns, and saving images

**Note:** Each component can be examined separately, added to, and deleted as necessary by the human to meet the intended goal.
